# Mediation analyses link cardiometabolic factors and liver fat with white matter hyperintensities and cognitive performance: A UK Biobank study

**DOI:** 10.1101/2024.10.17.24315664

**Authors:** Daniel E. Askeland-Gjerde, Lars T. Westlye, Patrik Andersson, Max Korbmacher, Ann-Marie de Lange, Dennis van der Meer, Olav B. Smeland, Sigrun Halvorsen, Ole A. Andreassen, Tiril P. Gurholt

**Author notes:** **Corresponding author:** Daniel E. Askeland-Gjerde, MD, Division of Mental Health and Addiction, Oslo University Hospital, Kirkeveien 166, 0450 Oslo (d.e.askeland-).

## Abstract

**Background:** Liver fat is associated with cardiometabolic disease, cerebrovascular disease, and dementia. Cerebrovascular disease, most frequently cerebral small vessel disease, identified by MRI as white matter hyperintensities (WMH), often contributes to dementia. However, liver fat’s role in the relationship between cardiometabolic risk, WMH, and cognitive performance is unclear.

**Methods:** In the UK Biobank cohort (n=32,628; 52.6% female; mean age 64.2±7.7 years; n=23,467 cognitive performance subsample), we used linear regression to investigate associations between cardiometabolic factors measured at baseline and liver fat, WMH, and cognitive performance measured at follow-up, on average, 9.3±2.0 years later. We used structural equation modeling to investigate whether liver fat mediates associations between cardiometabolic factors and WMH and whether WMH mediates associations between liver fat and cognitive performance.

**Results:** Nearly all cardiometabolic factors were significantly associated with liver fat (|r| in [0.03,0.41], p in [1.4x10^-8^,0〉) and WMH (|r| in [0.05,0.14], p in [1.5x10^-13^,2.7x10^-148^]) in regression models. Liver fat was associated with WMH (r=0.09,p=3x10^-64^) and cognitive performance (r=-0.03,p=1.5x10^-7^). Liver fat mediated the associations between cardiometabolic factors and WMH (|βmediation| in [0.01,0.03], pmediation in [5.7x10^-9^,0〉) and WMH mediated the associations between liver fat and cognitive performance (βmediation=-0.01,pmediation≍0).

**Conclusions:** Our findings indicate that liver fat mediates associations between cardiometabolic factors and WMH and that WMH mediates the association between liver fat and cognitive performance. This suggests that liver fat might be important for understanding the effects of cardiometabolic factors on cerebrovascular disease and cognitive function. Experimental studies are warranted to determine relevant targets for preventing vascular-driven cognitive impairment.

## Introduction

Metabolic dysfunction-associated steatotic liver disease (MASLD), formerly nonalcoholic fatty liver disease (NAFLD) (1), affects 30% of adults (2) and is associated with cerebrovascular disease (3,4) and dementia (5,6). Vascular pathology, most frequently cerebral small vessel disease (CSVD) (7,8), is implicated in 50-70% of dementia cases (9–11). As MASLD (2) and dementia (12) cases are expected to increase, it is vital to understand liver fat’s role in the early stages of CSVD-driven cognitive impairment, both to identify at-risk individuals and establish efficient preventive measures and treatment strategies.

MASLD is characterized by excessive fat accumulation in liver cells and one or more cardiometabolic abnormalities (13). As the diagnoses of MASLD and NAFLD are highly concordant (14–19), we use the term MASLD for both diagnoses. MASLD are associated with CSVD (20–23), identified as white matter hyperintensities (WMH) of presumed vascular origin on magnetic resonance imaging (MRI), albeit inconsistently (24–27), and prior studies have rarely used continuous liver fat measures. However, advances in rapid MRI allow for accurate liver fat quantification in large samples (28–30), facilitating novel large-scale research.

Cardiometabolic risk factors are associated with WMH (31–34) and MASLD (35–39), and both WMH and MASLD are associated with cognitive performance (40–48), though MASLD inconsistently (27,49–51). Furthermore, the associations might be sexually dimorphic as males have higher risk of MASLD (52) and CSVD (53) and females have a higher risk of dementia (54). Despite the observed associations between cardiometabolic factors, liver fat, WMH, and cognitive performance, to our knowledge no study has explicitly tested for the role of liver fat in the relationship between cardiometabolic factors, WMH, and cognitive performance. We hypothesized that (i) cardiometabolic risk factors are associated with WMH and that liver fat mediates these associations, (ii) liver fat is associated with cognitive performance and that WMH mediate this association, and (iii) there are sex-related differences in these associations.

## Methods and materials

### Participant sample

This observational study used UK Biobank data (access number 27412) and was approved by the Norwegian Regional Committees for Medical and Health Research Ethics. All participants gave informed consent and could withdraw their consent (opt-out list dated April 26^th^, 2023). We included participants with liver and brain MRI (n=41,760), excluded participants with a history of chronic liver disease (except MASLD), malignancies of the liver, biliary tract, or central nervous system, encephalitis, myelitis, stroke, traumatic brain injury, and neurodegenerative and demyelinating disorders (n=767; **Table S1**), and excluded participants who lacked data on sex, education, anthropometric measurements, blood pressure, serum measures, smoking status, or alcohol consumption (n=8,376). The included participants (n=32,628) attended the baseline assessment between April 2007 and October 2010 and the follow-up assessment between August 2014 and April 2022 (follow-up, mean, 9.3±2.0 [range, 4.3, 14.9] years). A subsample completed cognitive testing at follow-up (n=23,467; i.e., cognitive subsample).

### Liver and brain MRI

Liver and brain MRI were performed at 4 sites (Cheadle, Newcastle, Reading, and Bristol) using Siemens 1.5T MAGNETOM Aera and 3T Skyra scanners, respectively (55,56). AMRA Researcher (AMRA Medical AB) estimated liver fat percentage (57), and the University of Oxford’s Wellcome Centre for Integrative Neuroimaging estimated WMH volume with FSL BIANCA (58,59) and intracranial volume (ICV) with FreeSurfer (60).

### Demographic and clinical data

From the baseline assessment, we included sex, education, ethnicity, body mass index (BMI), waist circumference, systolic blood pressure (SBP), diastolic blood pressure (DBP), high-sensitivity c-reactive protein (CRP), glycated hemoglobin (HbA1c), high-density lipoprotein (HDL) cholesterol, low-density lipoprotein (LDL) cholesterol, total cholesterol, and triglycerides. From the follow-up assessment, we included age, site, alcohol consumption, smoking status, liver fat, WMH, ICV, numeric memory, fluid intelligence, trail making test B, matrix test, symbol digit substitution, tower rearranging, paired associate learning, and pairs matching (**Table S2**). Assigned sex was gathered from NHS registers. We categorized education into higher (college or university degree), intermediate (A levels, O levels, or equivalent), and lower education (otherwise) and smoking status as current, former, and never. We calculated pulse pressure by subtracting DBP from SBP and alcohol consumption by converting total weekly and monthly alcohol consumption into grams of alcohol per week.

We classified participants with probable hypertension (blood pressure ≥140/90mmHg or antihypertensive treatment), diabetes (HbA1c ≥48mmol/mol or antidiabetic treatment), or dyslipidemia (HDL cholesterol <1.03mmol/L, LDL cholesterol >4.13mmol/L, total cholesterol ≥6.20mmol/L, triglycerides >2.25mmol/L, or lipid-lowering treatment (61)) based on clinical measurements and reported medication use (**Table S3**) in nurse-led interviews at the baseline assessment.

We also classified participants with probable steatotic liver disease based on current or former diagnostic criteria (MASLD, NAFLD, metabolic dysfunction-associated fatty liver disease (MAFLD)) as follows. Probable NAFLD: liver fat ≥5% and alcohol consumption <20/30g/day (female/male) (62). Probable MASLD: probable NAFLD and ≥1: BMI ≥25kg/m^2^ or ≥23kg/m^2^ (Asian), waist circumference ≥80/94cm (female/male) or waist circumference ≥80/90cm (Asian, female/male), blood pressure ≥130/85mmHg or antihypertensive treatment, HbA1c ≥39mmol/mol or diabetes, triglycerides ≥1.7mmol/L or lipid-lowering treatment, and HDL cholesterol ≤1.3/1.0mmol/L (female/male) or lipid-lowering treatment (63). Probable MAFLD: liver fat ≥5%, and diabetes, BMI ≥25kg/m^2^ or ≥23kg/m2 (Asian), or ≥2: waist circumference ≥88/102cm (female/male) or waist circumference ≥80/90cm (Asian, female/male), blood pressure ≥130/85mmHg or antihypertensive treatment, HbA1c ≥39mmol/mol, triglycerides ≥1.7mmol/L or lipid-lowering treatment, HDL cholesterol <1.3/1.0mmol/L (female/male) or lipid-lowering treatment, and CRP >2mg/L (64).

### Cardiometabolic principal component analysis

We conducted principal component analysis (PCA) across 11 correlated cardiometabolic variables (**Figure S1**) to create uncorrelated composite measures of cardiometabolic risk. To stabilize variances (65), we log-transformed CRP and triglycerides (due to non-normal distributions; **Figures S2-S3**) and standardized the remaining variables (i.e., BMI, waist circumference, SBP, DBP, pulse pressure, HbA1c, HDL cholesterol, LDL cholesterol, and total cholesterol) by mean centering and dividing by the standard deviation. As PCA is sensitive to outliers (65), we removed values deviating >3 standard deviations from the mean (**Table S4A**). 2,368 participants had ≥1 missing data point after outlier removal, but all participants had >50% of data points and were included in the subsequent analyses. Next, we imputed missing data points with the *missMDA* R-package (66), conducted the PCA with the *prcomp* R-function, and extracted the first 3 principal components (PCs) based on their explained variance, 31.9%, 23.6%, and 17.9%, respectively (**Figure S4**). PC1’s largest loadings were from SBP, DBP, and anthropometric measures (loadings in [0.37, 0.48]; **Figure S5**); PC2’s from cholesterol and anthropometric measures (|loadings| in [0.19, 0.64]); and PC3’s from pulse pressure, SBP, and anthropometric measures (|loadings| in [0.35, 0.50]). All PC1’s loadings indicated higher cardiometabolic risk.

### Cognitive principal component analysis

Vascular cognitive impairment can affect multiple cognitive domains (11). Therefore, we computed a measure of general cognitive performance using PCA across 8 cognitive tests from the follow-up assessment (**Figure S6**). We log-transformed trail making test B and pairs matching (due to non-normal distributions; **Figures S7-S8**) and standardized the remaining variables (i.e., numeric memory, fluid intelligence, matrix test, symbol digit substitution, tower rearranging, and paired associate learning). We did not include reaction time (data field 20023) as it might be lower correlated with general cognitive performance (67) and have a different genetic basis (68) and prospective memory (data field 20018) as it was a binary variable. We conducted the PCA as described above. After outlier removal (**Table S4B**), 11,579 participants had ≥1 missing data point. 23,467 participants had >50% data points and were included in subsequent analyses. We imputed missing data points, computed the PCA, and extracted PC1 based on its explained variance (40.7%; **Figure S9**). All cognitive tests contributed to PC1, with largest contributions from fluid intelligence, matrix test, and tower rearranging (loadings in [0.40, 0.46]; **Figure S10**). All PC1’s loadings indicated better cognitive task performance, suggesting that cognitive PC1 is a measure of general cognitive performance.

### Statistical analysis

We used R version 4.2.0 (69) and R-packages *tidyverse* (70), *forcats* (71), *ggplot2* (72), *patchwork* (73), and *lavaan* (74) for analyses and data visualization. Continuous variables were assessed using histograms and quantile-quantile plots (**Figures S2-S3, S7-S8**). As residuals from linear regression analyses with liver fat, WMH, trail making test B, and pairs matching as outcomes and CRP and triglycerides as predictors were non-normal, we log-transformed these variables. Remaining continuous variables were standardized by mean-centering and dividing by the standard deviation. We used the total sample (n=32,628) for the analyses with liver fat and WMH as outcomes and the cognitive subsample (n=23,467) for the analyses with the cognitive outcome measures. We tested for sex-related differences in the total sample (**Table S5**) and differences between the total sample and cognitive subsample (**Table S6**) using t-tests and chi-squared tests.

First, we conducted multiple linear regression analyses with liver fat, WMH, and cognitive PC1 as outcomes to verify the assumptions of our planned mediation analyses. We used the cardiometabolic risk factors and cardiometabolic PC1-PC3 as predictors for all outcomes, liver fat and probable steatotic liver disease as predictors for WMH and cognitive PC1, and WMH as a predictor for cognitive PC1. Each predictor was analyzed by itself in a separate model. Next, we performed analyses with an interaction term between the predictor and sex to test for sex-related differences. For analyses with cognitive PC1 as the outcome, we conducted follow-up analyses with individual cognitive tests. The regression analyses were adjusted for age, age^2^, sex, age-by-sex, age^2^-by-sex, site, smoking status, alcohol consumption, ICV (WMH only), and education (cognitive analyses only).

Next, we performed SEM mediation analyses (**Figure 1**) using the *sem function* in the *lavaan R-package* version 0.6-11 (74). We computed standard errors with bootstrapping using 10,000 draws. Each model consists of two regression equations and includes outcome (*Y*), mediator (*M*), predictor (*X*), intercepts (*i*), and error terms (*e*). Additionally, we performed sex-stratified analyses for predictors that had significant sex interactions in relevant regression analyses. We did not include interaction terms as *lavaan* does not support them (74).

**Figure 1:**
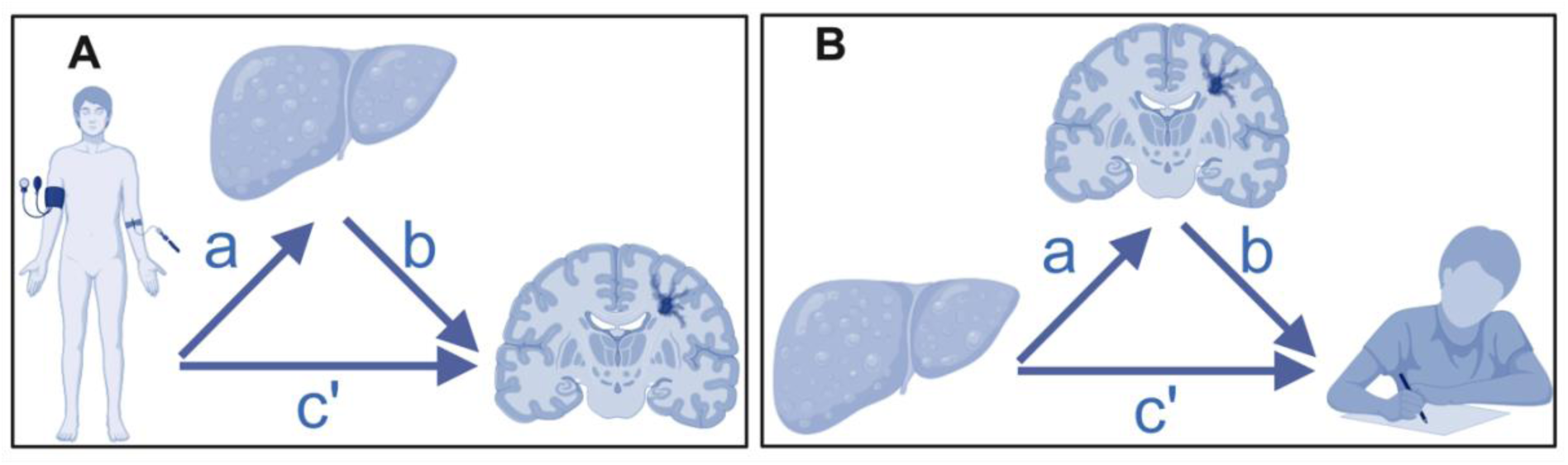
Path diagram of the mediation analyses. The figure shows the path diagrams of the mediation analyses with **A** white matter hyperintensities as outcome, liver fat as mediator, and cardiometabolic factors as predictors and with **B** cognitive principal component 1 as outcome, white matter hyperintensities as mediator, and liver fat and steatotic liver disease as predictors. (Created with BioRender.com.)

In the first set of mediation analyses, we used WMH as outcome, liver fat as mediator, cardiometabolic factors and cardiometabolic PC1-PC3 separately as predictors (**Figure 1A**) using the following model:

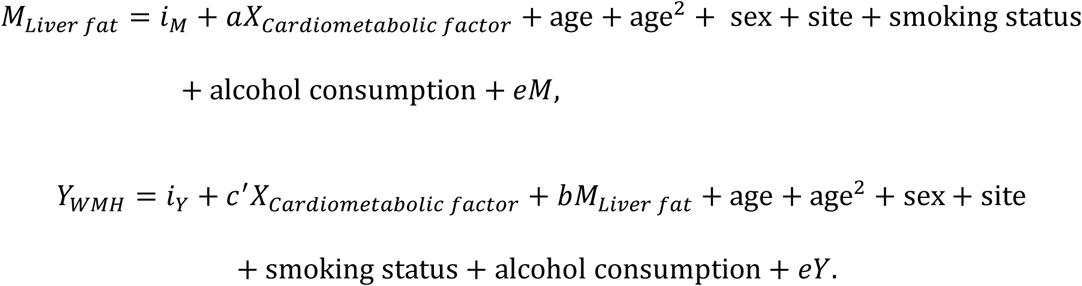

Initially, we included ICV as a covariate in the second equation. However, model fit measures were poor. After removing ICV from the equation, the models were saturated and had good model fit measures: confirmatory fit index (CFI) in [1.000, 1.000], Tucker-Lewis Index (TLI) in [1.000, 1.000], root mean square error of approximation (RMSEA) in [0.000, 0.000], and standardized root mean squared residual (SRMR) in [1.0x10^-16^, 9.8x10^-18^].

In the second set of mediation analyses, we used cognitive PC1 as outcome, WMH as mediator, and liver fat and probable steatotic liver disease separately as predictors (**Figure 1B**) using the following model:

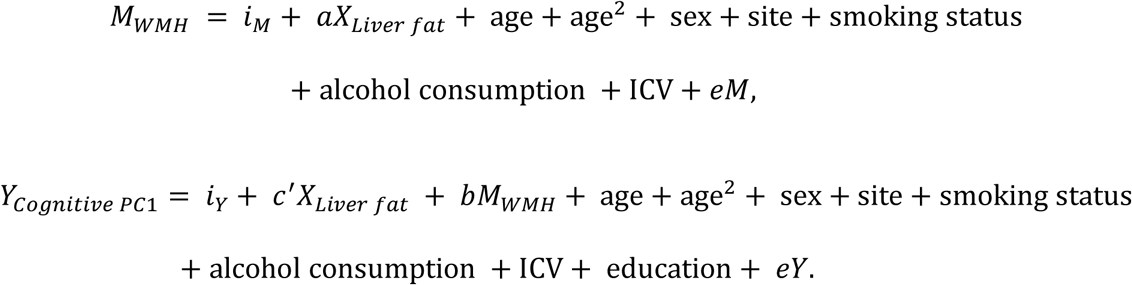

The fit measures were good: CFI in [0.993, 1.000], TLI in [0.918, 1.000], RMSEA in [0.034, 0.000], and SRMR in [0.001, 0.005]. We conducted equivalent follow-up mediation analyses on individual cognitive tests, with good fit measures: CFI in [0.998, 1.000], TLI in [0.975, 0.998], RMSEA in [0.007, 0.025], and SRMR in [0.002, 0.004].

We report partial correlation coefficients (r) (75) from the linear regression analyses and standardized regression coefficients (β) for direct, indirect, and total effects from the mediation analyses. We derived a study-wide Bonferroni threshold p≤0.05/N=0.05/156=0.0003, where N is the number of analyses. We planned 28 (14 sex-specific), 36 (18 sex-specific), and 38 (19 sex-specific) regression analyses with liver fat, WMH, and cognitive PC1 as outcomes, respectively, and 42 (28 sex-specific) and 12 (8 sex-specific) mediation analyses with WMH and cognitive PC1 as outcomes, respectively. Follow-up analyses on individual cognitive tests were not included in the analysis count. Results are described as significant if they pass the Bonferroni threshold and we report unadjusted p-values. Since R uses double-precision values, some p-values are reported as 0, which indicate approximately equal to 0.

## Results

### Sample description

The sample consisted of 32,628 UK Biobank participants (n=17,164 (52.6%) females; **Table S5**), who were mostly middle- to late-aged (mean age 64.2±7.7 years; range [48, 83]) and had completed higher (46.5%) or intermediate education (32.7%). On average, males had higher BMI (27.08±3.64 vs. 26.03±4.52) and liver fat (4.70±4.18 vs. 3.86±3.97), and higher risk of probable hypertension (53.5% vs. 36.1%), diabetes (3.1% vs. 1.5%), and dyslipidemia (61.0% vs. 46.2%). Probable steatotic liver disease prevalences varied based on the diagnostic criteria used, but males had consistently higher risk of steatotic liver diseases, probable NAFLD (20.3% vs. 15.5%), MASLD (19.8% vs. 15.0%), and MAFLD (26.8% vs. 17.0%). The cognitive subsample (n=23,467) had comparable demographic, clinical, and imaging data as the total sample (**Table S6**).

### Cardiometabolic factors and liver fat

Multiple linear regression analyses revealed significant associations between all cardiometabolic factors and liver fat (**Figure 2AB, Table S7**). BMI, waist circumference, triglycerides, and cardiometabolic PC1 (largest loadings from anthropometric and blood pressure measurements) showed medium-to-large effects (r in [0.323, 0.407], p-values ≍ 0). The remaining variables showed small-to-medium effects (|r| in [0.031, 0.269], p-values in [1.4x10^-8^, 8.9x10^-246^]). All variables except HDL cholesterol and PC3 (negative loadings from BMI and waist circumference) were associated with higher liver fat.

**Figure 2:**
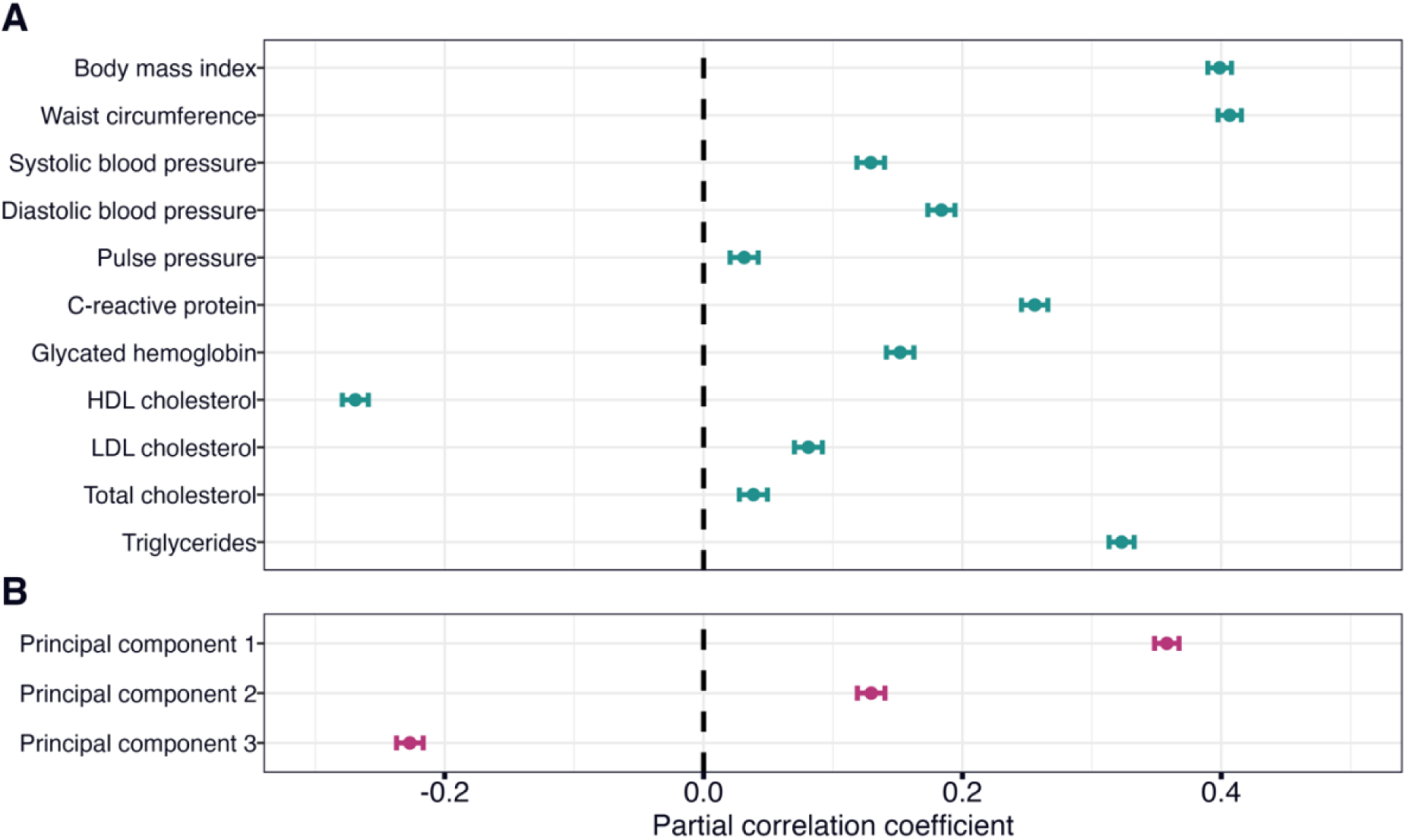
Linear associations between cardiometabolic factors and liver fat. The figure shows forest plots with the associations of **A** cardiometabolic risk factors and **B** cardiometabolic principal components with liver fat. The error bars correspond to 95% confidence intervals. The regression models were adjusted for age, age^2^, sex, age-by-sex, age^2^-by-sex, site, smoking status, and alcohol consumption. HDL, high-density lipoprotein; LDL, low-density lipoprotein.

Multiple linear regression analyses revealed significant interactions between sex and BMI, DBP, CRP, LDL cholesterol, and triglycerides on liver fat (**Table S8**). An increase in BMI was associated with a steeper increase in liver fat in males than females (r=0.028, p=4.5x10^-7^), while increases in DBP, CRP, LDL cholesterol, and triglycerides were associated with a steeper increase in liver fat in females than males (r in [-0.024, -0.070], p-values in [2.2x10^-5^, 7.7x10^-37^]).

### Cardiometabolic factors, liver fat, and white matter hyperintensities

Linear regression revealed significant associations between all cardiometabolic risk factors (except LDL cholesterol, total cholesterol, and cardiometabolic PC3) and WMH (**Figure 3AB, Table S9**) with small-to-medium effects (r in [0.041, 0.143], p-values in [1.5x10^-13^, 2.7x10^-148^]). BMI, waist circumference, SBP, DBP, and cardiometabolic PC1 had the largest effects. Liver fat and probable steatotic liver diseases were associated with higher WMH volume with small effects (r in [0.066, 0.094], p-values in [9.6x10^-33^, 3.0x10^-64^]; **Figure 3C**).

**Figure 3:**
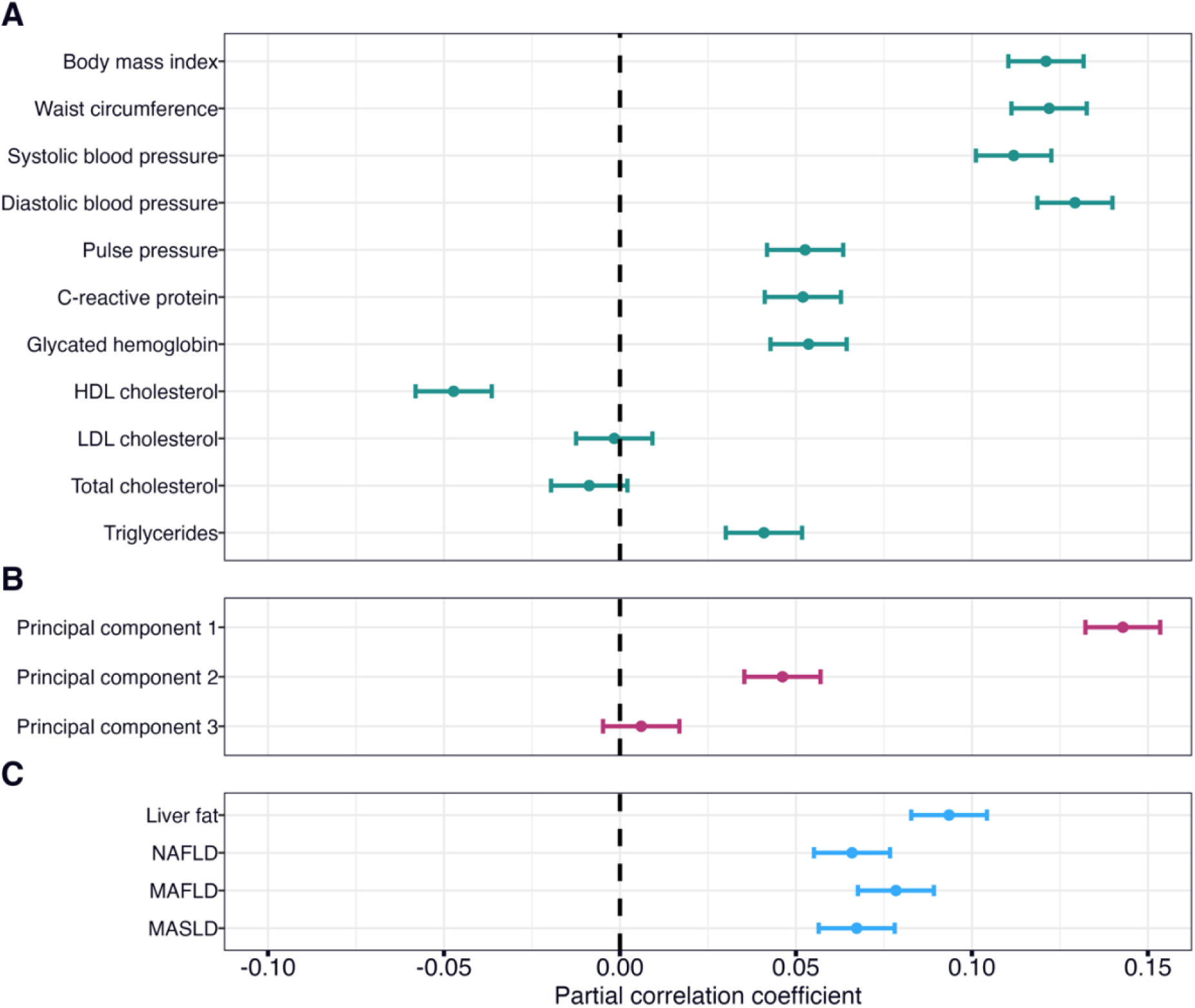
Linear associations between cardiometabolic factors, liver fat, and white matter hyperintensities. The figure shows forest plots with the associations of **A** cardiometabolic risk factors, **B** cardiometabolic principal components, and **C** liver fat and probable steatotic liver disease with white matter hyperintensities. The error bars correspond to 95% confidence intervals. The regression models were adjusted for age, age^2^, sex, age-by-sex, age^2^-by-sex, site, smoking status, alcohol consumption, and intracranial volume. HDL, high-density lipoprotein; LDL, low-density lipoprotein; NAFLD, nonalcoholic fatty liver disease; MAFLD, metabolic dysfunction-associated fatty liver disease; MASLD, metabolic dysfunction-associated steatotic liver disease.

Linear regression analyses revealed significant interactions between sex and BMI, waist circumference, and cardiometabolic PC2 on WMH (**Table S10**). An increase in BMI, waist circumference, and cardiometabolic PC2 (largest loadings from BMI and waist circumference) was associated with a steeper increase in WMH in males than females (r in [0.022, 0.047], p- values in [7.2x10^-5^, 3.3x10^-17^]).

### Cardiometabolic factors, liver fat, white matter hyperintensities, and cognitive performance

Linear regression revealed significant associations between cognitive PC1 and BMI, SBP, DBP, HbA1c, and cardiometabolic PC1 (r in [-0.027, -0.037], p-values in [3.9x10^-5^, 1.2x10^-8^]; **Figure 4AB, Table S11**). Liver fat and probable steatotic liver diseases were significantly associated with cognitive PC1 (r in [-0.030, -0.034], p-values in [3.5x10^-6^, 1.5x10^-7^]; **Figure 4C**). Of all variables, WMH had the largest effect on cognitive PC1 (r=-0.071, p=1.2x10^-27^; **Figure 4D**). None of the predictors had significant interactions with sex (**Table S12**).

**Figure 4:**
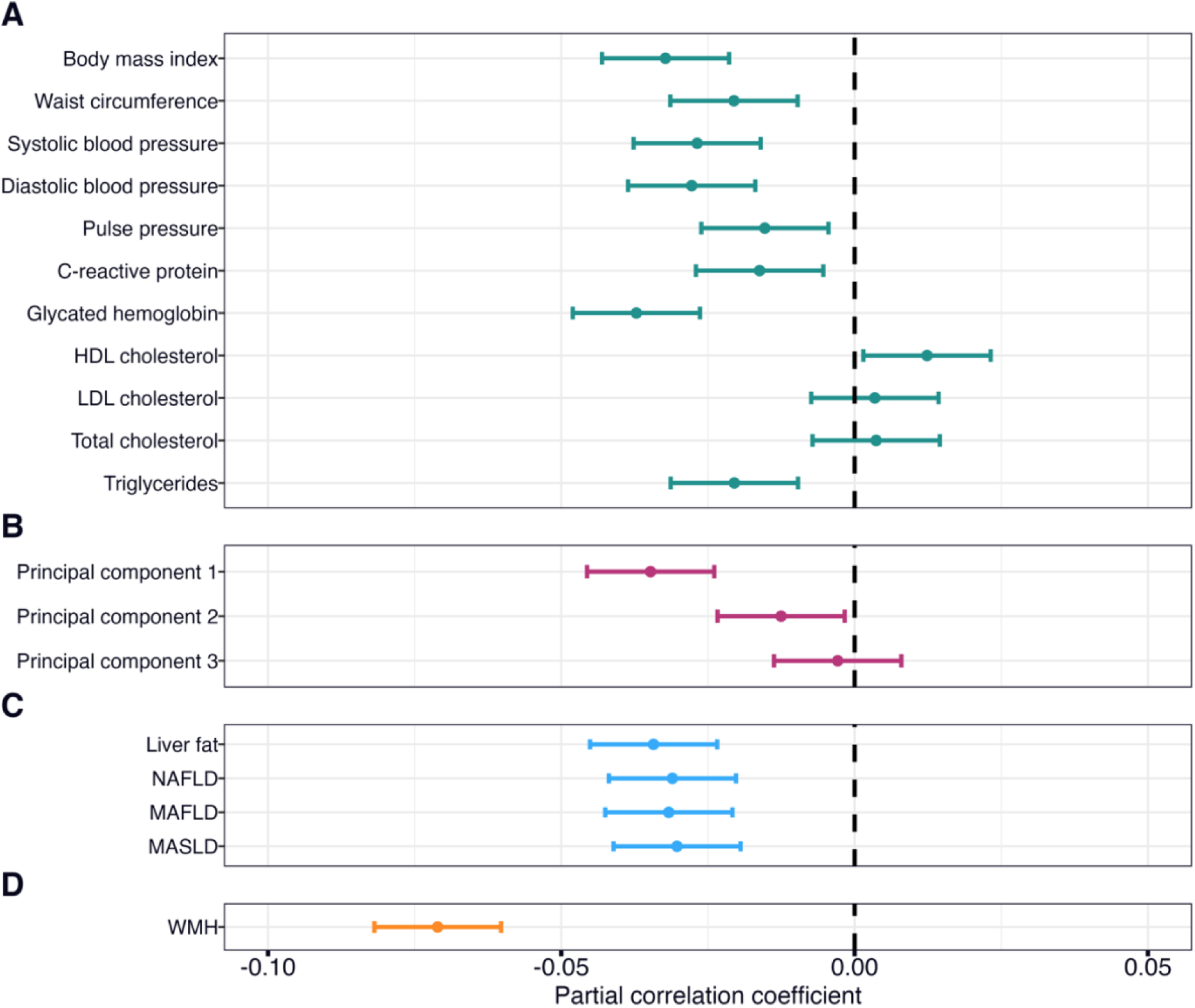
Linear associations between cardiometabolic factors, liver fat, WMH, and the cognitive principal component 1. The figure shows forest plots with the associations of **A** cardiometabolic risk factors, **B** cardiometabolic principal components, **C** liver fat and probable steatotic liver disease, and **D** white matter hyperintensities with cognitive principal component 1. The error bars correspond to 95% confidence intervals. The regression models were adjusted for age, age^2^, sex, age-by-sex, age^2^-by-sex, site, smoking status, alcohol consumption, education, and intracranial volume (only white matter hyperintensities). HDL, high-density lipoprotein; LDL, low-density lipoprotein; NAFLD, nonalcoholic fatty liver disease; MAFLD, metabolic dysfunction-associated fatty liver disease; MASLD, metabolic dysfunction-associated steatotic liver disease; WMH, white matter hyperintensities.

In follow-up analyses on individual cognitive tests, cardiometabolic factors were significantly associated with a range of cognitive tests, most often numeric memory, matrix test, and paired associate learning (**Table S13**). Liver fat was significantly associated with all cognitive tests, except trail making test B and pairs matching. There were no significant interactions between any predictors and sex (**Table S14**).

### Liver fat mediates the associations between cardiometabolic factors and white matter hyperintensities

SEM mediation analyses revealed significant total and mediation (i.e., indirect) effects via liver fat on WMH for all cardiometabolic factors except LDL cholesterol, total cholesterol, and cardiometabolic PC3 (**Table S15**). BMI, waist circumference, SBP, and DBP had the largest direct effects (β in [0.086, 0.122], p-values ≍ 0; **Figure 5AC**), while waist circumference, HDL cholesterol, and triglycerides had the largest mediation effects (|β| in [0.020, 0.026], p-values ≍ 0; **Figure 5BD**). The direct effects of HDL cholesterol and triglycerides were not significant, indicating that their associations with liver fat fully explain their associations with WMH.

**Figure 5:**
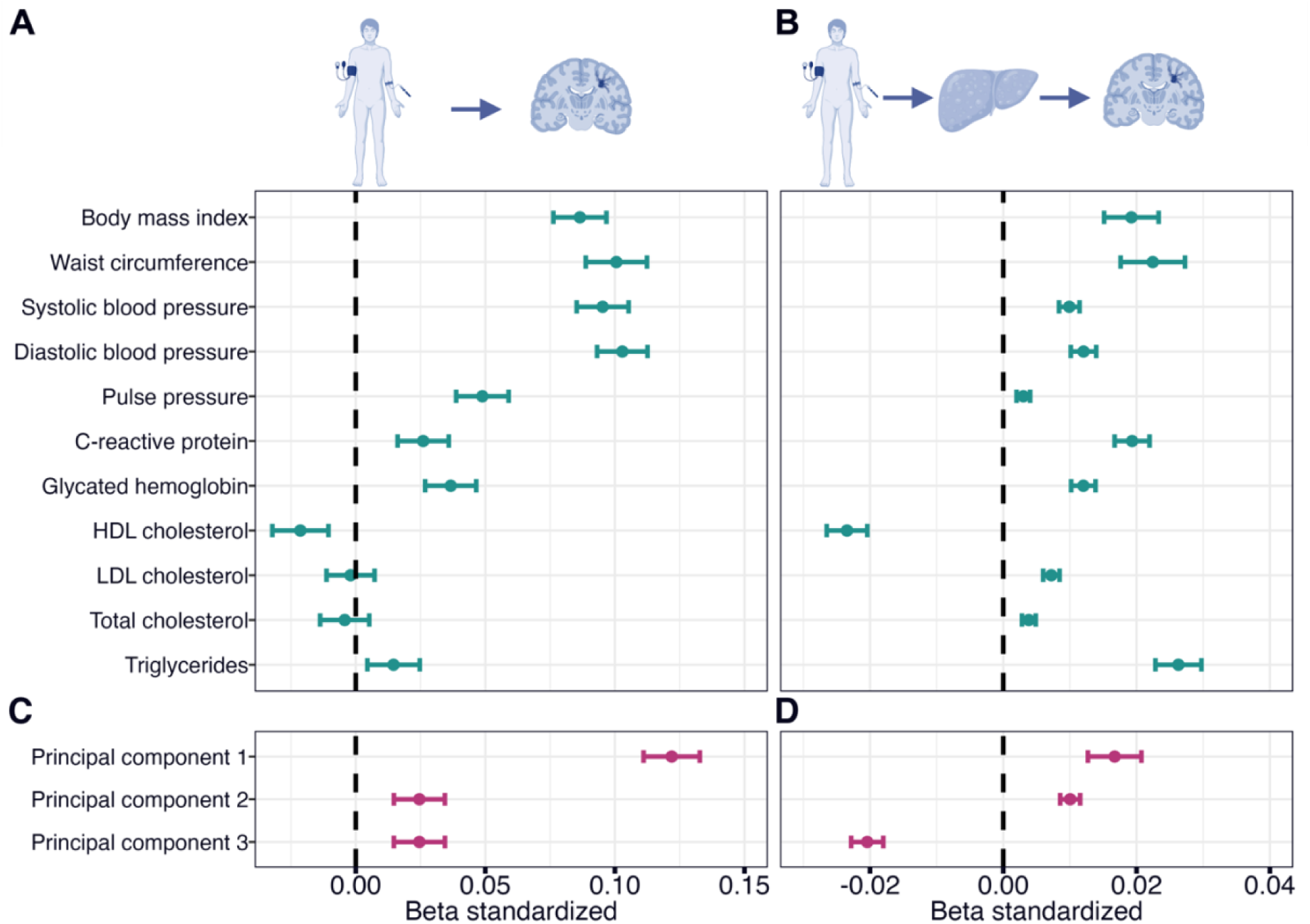
Mediation analyses with white matter hyperintensities as the outcome and liver fat as the mediator. The figure shows forest plots with the **A** direct and **B** mediation (i.e, indirect) effects via liver fat of cardiometabolic risk factors and the **C** direct and **D** mediation effects via liver fat of cardiometabolic principal components on white matter hyperintensities. Error bars correspond to standardized 95% confidence intervals. HDL, high-density lipoprotein; LDL, low-density lipoprotein. Illustrations created with BioRender.com.

### Liver fat mediates the associations between cardiometabolic factors and white matter hyperintensities: Sex-related differences

Sex-stratified analyses (**Table S16**) revealed higher direct effects (**Figure 6A**) on WMH of BMI and waist circumference in males (β in [0.11, 0.12], p-values ≍ 0) than females (β=0.07, p-values ≍ 0), while the mediation effects via liver fat (**Figure 6B**) were similar (β=0.02, p-values in [3.9x10^-8^, 2.2x10^-16^]). Furthermore, cardiometabolic PC2 (largest loadings from anthropometric and cholesterol measures) had a significant total effect on WMH in males (β=0.06, p=2.3x10^-10^) but not in females (β=0.02, p=0.001). The results indicate that the direct effects of anthropometric measures on WMH might be stronger in males than females, while the mediation effects via liver fat are similar.

**Figure 6:**
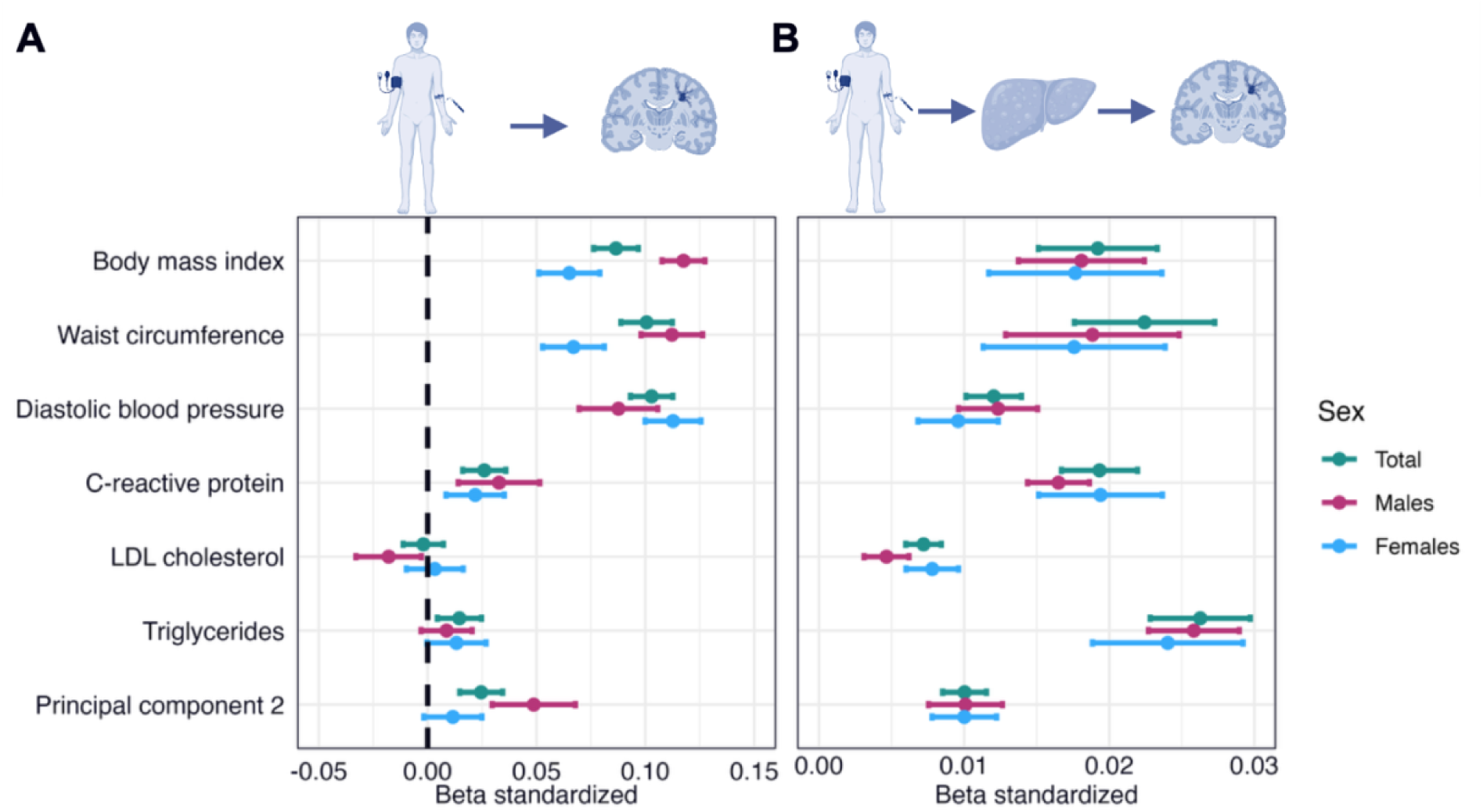
Mediation analyses with white matter hyperintensities as the outcome and liver fat as the mediator in the total sample and in males and females separately. The figure shows forest plots with the **A** direct and **B** mediation effects via liver fat of cardiometabolic risk factors and the **C** direct and **D** mediation effects via liver fat of cardiometabolic principal components on white matter hyperintensities. Error bars correspond to standardized 95% confidence intervals. HDL, high-density lipoprotein; LDL, low-density lipoprotein. Illustrations created with BioRender.com.

### White matter hyperintensities mediate the associations of liver fat and probable steatotic liver diseases with cognitive performance

SEM mediation analyses revealed significant total and mediation effects via WMH on cognitive PC1 for both liver fat and probable steatotic liver diseases (**Table S17**). Effect sizes were comparable across predictors, for direct (β in [-0.020, -0.021], p-values in [0.0008, 0.0003]; **Figure 7A**) and mediation (β in [-0.004, -0.006], p-values in [4x10^-14^, 0〉; **Figure 7B**) effects.

**Figure 7:**
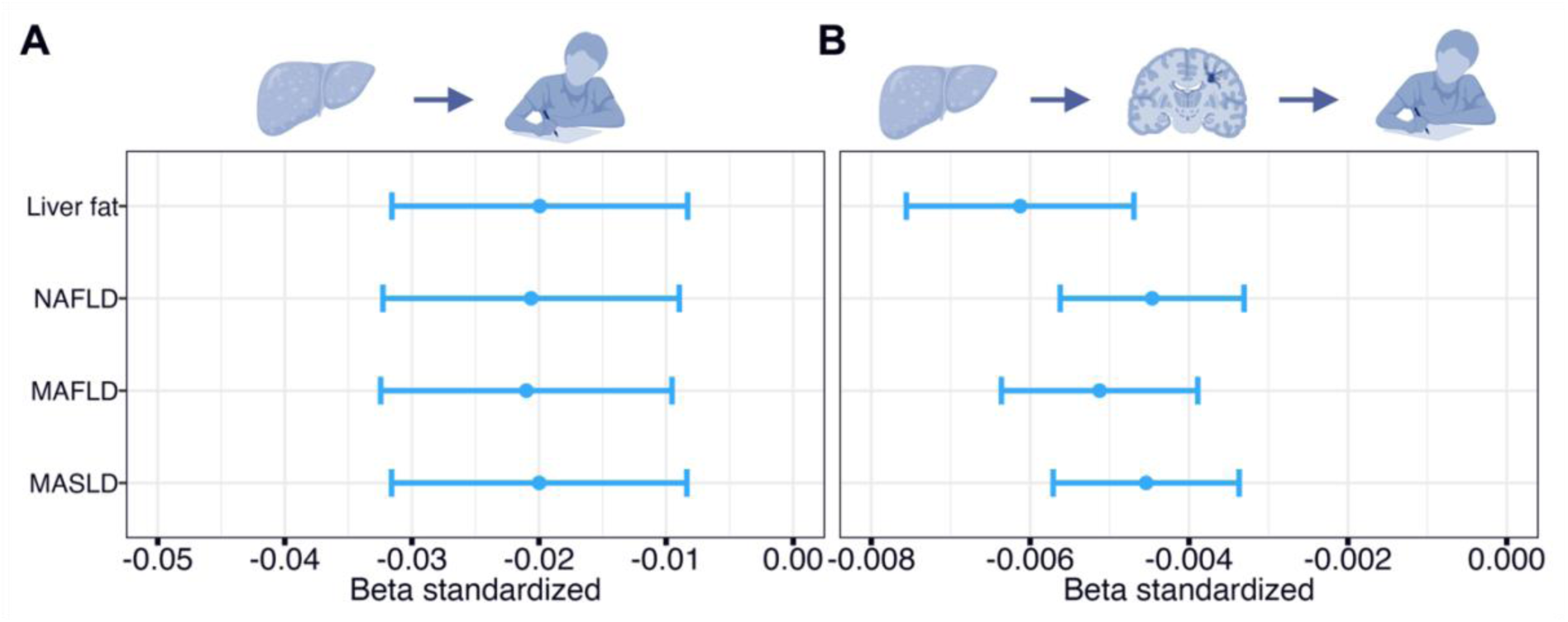
Mediation analyses with cognitive principal component 1 as the outcome and white matter hyperintensities as the mediator. The figure shows forest plots with the **A** direct and **B** mediation effects via WMH of liver fat and probable steatotic liver disease on cognitive principal component 1. Error bars correspond to standardized 95% confidence intervals. NAFLD, nonalcoholic fatty liver disease; MAFLD, metabolic dysfunction-associated fatty liver disease; MASLD, metabolic dysfunction-associated steatotic liver disease. Illustrations created with BioRender.com.

In follow-up analyses on individual cognitive tests, WMH mediated the associations between liver fat and numeric memory, symbol digit substitution, and paired associate learning (**Table S18**).

## Discussion

In this study, we showed that liver fat mediates the associations between cardiometabolic risk factors and higher WMH and that WMH mediates the associations between higher liver fat and lower cognitive performance in middle- to late-aged participants. Furthermore, higher BMI and waist circumference might be more strongly associated with liver fat and WMH in males than in females. Our results implicate liver fat in CSVD and cognitive performance and indicate that a higher burden of WMH might partly explain the link between higher liver fat and lower cognitive performance. Liver fat might, therefore, be a relevant treatment target to prevent the development of vascular cognitive impairment.

SEM mediation analyses revealed that liver fat mediates the link between nearly all cardiometabolic factors and WMH, expanding our results from the corresponding regression analyses. Our findings align with observed associations between WMH and BMI (34), blood pressure (76–78), CRP (79), HbA1c (80,81), HDL cholesterol (82), and triglycerides (83). The link between liver fat and WMH might be explained by liver fat’s association with exacerbated cardiometabolic risk (84–87) and inflammatory factors such as homocysteine (88–91). While causal interpretations remain speculative, our results implicate liver fat in the link between cardiometabolic risk and higher WMH volume.

The cardiometabolic factors BMI, waist circumference, CRP, HDL cholesterol, triglycerides, and cardiometabolic PC1 (largest loadings from anthropometric and blood pressure measures) had the overall largest mediation effects via liver fat on WMH. Additionally, the associations of HDL cholesterol and triglycerides with liver fat fully explained their associations with WMH. Our findings align with the strong links between liver fat and anthropometric measures (36) and the hypothesis that ectopic fat (e.g., liver fat) might be more strongly associated with cerebrovascular disease than subcutaneous fat (3,4,92), and observations that liver fat might initiate inflammatory pathways (93–95), alter lipid and lipoprotein regulation (87), and contribute to higher blood pressure through higher vasoconstriction (96) and impaired peripheral vasodilation (97). Taken together, our findings might suggest that interventions aimed at general and abdominal obesity, dyslipidemia, and low-grade inflammation, could be particularly beneficial in preventing liver fat accumulation and WMH development.

In the current sample, males had higher cardiometabolic risk, liver fat, and WMH volume on average than females. Regression analyses revealed steeper increases in liver fat and WMH per increase in BMI in males, aligning with epidemiological findings in steatotic liver disease (98,99) and CSVD (53). In SEM mediation analyses, the cardiometabolic PC2, with loadings from BMI and waist circumference, was only significantly associated with WMH in males, and the direct effect sizes of BMI and waist circumference were larger in males than females, while indirect effects via liver fat were similar. We might speculate that males store less fat in the subcutis than females (100–102), leading to a harmful body fat distribution that could contribute to CSVD and possibly other brain outcomes, as similar patterns have been shown for brain age (103,104).

We show that WMH mediates the associations between lower general cognitive performance and liver fat and probable steatotic liver disease, expanding on previous observations on associations between MASLD and WMH (20–23) and cognitive performance (40–48). In follow-up analyses, we found that WMH mediates the associations between liver fat and lower performance on numeric memory, symbol digit substitution, and paired associate learning tests. These tests all contributed to the cognitive PC1 (i.e., general cognitive performance) and cover working memory, processing speed, and verbal declarative memory (67), cognitive domains often affected by vascular cognitive impairment (11). Our findings might suggest a role of liver fat in the development of CSVD-driven cognitive decline. Improving cardiometabolic health is one of the strategies outlined to prevent dementia cases (105), and our findings suggest that lowering liver fat might also be relevant, as previously shown for thigh muscle-fat-infiltration (106). Importantly, our study demonstrates the close links between general cardiometabolic risk and liver fat. Weight loss interventions can be effective in lowering liver fat (107). However, weight loss does not always lead to MASLD improvement, especially in severe cases (108,109). Therefore, interventions for preventing liver fat accumulation in the general population are needed.

Our study has strengths and limitations. It is significantly larger than previous studies and assesses liver fat (57) and WMH (59) with accurate, quantitative methods. We used a well-characterized sample, individual and composite cardiometabolic factors, liver fat percentage and steatotic liver disease diagnoses, general cognitive performance and individual cognitive tests, and tested for sex differences. However, we only assessed WMH as it was beyond the scope of this paper to investigate other CSVD markers. UK Biobank participants are healthier, wealthier, and less ethnically diverse than the general UK population (110,111), which might limit the generalizability of our findings. Although brain MRI and cognitive testing were performed years after cardiometabolic assessment, we did not use imaging data from multiple time points. Therefore, we cannot fully exclude different directions of effects, and differently designed studies are needed to make causal claims.

## Conclusion

Our findings suggest that liver fat might play a role in CSVD both directly and by mediating the associations between cardiometabolic risk factors and higher WMH volume. Higher BMI and waist circumference might be more strongly associated with liver fat and WMH in males than in females, while the link between liver fat and WMH appears to be similar in both sexes. Our results indicate links between liver fat and cognitive performance, both for general cognitive performance, working memory, processing speed, and verbal declarative memory. The associations with cognitive performance were mediated by higher WMH volume, suggesting that liver fat could contribute to the development of vascular cognitive impairment. Our findings warrant experimental studies on the underlying mechanisms and on liver fat as a potential target for preventing or delaying cognitive decline.

## Data availability

The R-code used for this project is freely available on https://github.com/deagjerde/cardiometabolic-brain-link-via-liver-mediation.

## Acknowledgments

This research has been conducted using the UK Biobank Resource under Application Number 27412. This work uses data provided by patients and collected by the NHS as part of their care and support. We performed all data analyses on the Services for Sensitive Data (TSD), University of Oslo, Norway, with resources from UNINETT Sigma2 – the National Infrastructure for High-Performance Computing and Data Storage in Norway.

## Authors contribution

**DEAG:** Conceptualization, Software, Formal analysis, Data curation, Writing – Original draft, Visualization. **LTW:** Resources, Writing – Review & editing, Project administration, Funding acquisition. **PA:** Resources, Writing – Review & editing. **MK:** Writing – Review & editing. **AMDL:** Writing – Review & editing, Funding acquisition. **DVDM:** Writing – Review & editing. **OBS:** Writing – Review & editing. **SH:** Conceptualization, Writing – Review & editing, Supervision. **OAA:** Conceptualization, Resources, Writing – Review & editing, Supervision, Project administration, Funding acquisition. **TPG:** Conceptualization, Software, Formal analysis, Writing – Review & editing, Visualization, Supervision, Project administration, Funding acquisition. All authors revised the manuscript and approved the final version.

