## Supplementary Materials for "Mediation analyses link cardiometabolic factors and liver fat with white matter hyperintensities and cognitive performance: A UK Biobank study"

**Affiliations:**
[1] Section for Precision Psychiatry, Division of Mental Health and Addiction, Oslo University Hospital, Oslo, Norway
[2] Institute of Clinical Medicine, University of Oslo, Oslo, Norway
[3] Department of Psychology, University of Oslo, Oslo, Norway
[4] KG Jebsen Centre for Neurodevelopmental Disorders, University of Oslo, Oslo, Norway
[5] AMRA Medical AB, 58222 Linköping, Sweden
[6] Neuro-SysMed Center of Excellence for Clinical Research in Neurological Diseases, Department of Neurology, Haukeland University Hospital, Bergen, Norway
[7] Mohn Medical Imaging and Visualization Centre (MMIV), Department of Radiology, Haukeland University Hospital, Bergen, Norway
[8] Department of Health and Functioning, Western Norway University of Applied Sciences, Bergen, Norway
[9] Department of Clinical Neurosciences, Lausanne University Hospital (CHUV) and University of Lausanne, Lausanne, Switzerland
[10] Department of Psychiatry, University of Oxford, Oxford, UK
[11] School of Mental Health and Neuroscience, Faculty of Health, Medicine and Life Sciences, Maastricht University, Maastricht, The Netherlands
[12] Department of Cardiology, Oslo University Hospital and University of Oslo, Oslo, Norway

### Supplementary Tables

#### Table S1: ICD-10 diagnoses of exclusion

| Code | Name | Code | Name | Code | Name |
|---|---|---|---|---|---|
| C22 | Malignant neoplasm of liver and intrahepatic bile ducts | G11 | Hereditary ataxia | I63 | Cerebral infarction |
| C23 | Malignant neoplasm of gallbladder | G12 | Spinal muscular atrophy and related syndromes | I64 | Stroke, not specified as hemorrhage or infarction |
| C24 | Malignant neoplasm of other and unspecified parts of biliary tract | G13 | Systemic atrophies primarily affecting central nervous system in diseases classified elsewhere | I69 | Sequelae of cerebrovascular disease |
| C70 | Malignant neoplasm of meninges | G14 | Postpolio syndrome | K70 | Alcoholic liver disease |
| C71 | Malignant neoplasm of brain | G20 | Parkinson's disease | K71 | Toxic liver disease |
| C72 | Malignant neoplasm of spinal cord, cranial nerves and other parts of central nervous system | G23 | Other degenerative diseases of basal ganglia | K72 | Hepatic failure, not elsewhere classified |
| F00 | Dementia in Alzheimer's disease | G30 | Alzheimer's disease | K73 | Chronic hepatitis, not elsewhere classified |
| F01 | Vascular dementia | G31 | Other degenerative diseases of nervous system, not elsewhere classified | K74 | Fibrosis and cirrhosis of liver |
| F02 | Dementia in other diseases classified elsewhere | G32 | Other degenerative disorders of nervous system in diseases classified elsewhere | K75 | Other inflammatory liver diseases |
| F03 | Unspecified dementia | G35 | Multiple sclerosis | S06.2 | Diffuse brain injury |
| F06.7 | Mild cognitive disorder | G36 | Other acute disseminated demyelination | S06.3 | Focal brain injury |
| G04 | Encephalitis, myelitis and encephalomyelitis | G37 | Other demyelinating diseases of central nervous system | S06.4 | Epidural hemorrhage |
| G05 | Encephalitis, myelitis and encephalomyelitis in diseases classified elsewhere | I60 | Subarachnoid hemorrhage | S06.5 | Traumatic subdural hemorrhage |
| G06 | Intracranial and intraspinal abscess and granuloma | I61 | Intracerebral hemorrhage | S06.6 | Traumatic subarachnoid hemorrhage |
| G10 | Huntington's disease | I62 | Other nontraumatic intracranial hemorrhage | S06.7 | Intracranial injury with prolonged coma |

**Table S2: Included UK Biobank data fields**

| ID | Name | ID | Name | ID | Name |
|---|---|---|---|---|---|
| 41280 | Date of first in-patient diagnosis - ICD10 | 41270 | Diagnoses - ICD10 | 30870 | Triglycerides |
| 30780 | LDL direct | 30760 | HDL cholesterol | 30750 | Glycated hemoglobin (HbA1c) |
| 30710 | C-reactive protein | 30690 | Cholesterol | 26521 | Estimated intracranial volume |
| 25781 | Total volume of white matter hyperintensities (from T1 and T2_FLAIR images) | 24352 | FR liver PDFF mean | 23324 | Number of symbol digit matches made correctly |
| 22436 | Liver PDFF (proton density fat fraction) | 21004 | Number of puzzles correct | 21003 | Age when attended assessment center |
| 21001 | Body mass index (BMI) | 21000 | Ethnic background | 20197 | Number of word pairs correctly associated |
| 20139 | Number of letters correctly identified | 20117 | Alcohol drinker status | 20116 | Smoking status |
| 20023 | Mean time to correctly identify matches | 20016 | Fluid intelligence score | 20003 | Treatment/medication code |
| 10722 | Qualifications (pilot) | 10137 | Number of incorrect matches in round (pilot) | 6373 | Number of puzzles correctly solved |
| 6350 | Duration to complete alphanumeric path | 6348 | Duration to complete numeric path | 6138 | Qualifications |
| 5364 | Average weekly intake of other alcoholic drinks | 4462 | Average monthly intake of other alcoholic drinks | 4451 | Average monthly fortified wine intake |
| 4440 | Average monthly spirits intake | 4429 | Average monthly beer plus cider intake | 4418 | Average monthly champagne plus white wine intake |
| 4407 | Average monthly red wine intake | 4282 | Maximum digits remembered correctly | 4080 | Systolic blood pressure, automated reading |
| 4079 | Diastolic blood pressure, automated reading | 1608 | Average weekly fortified wine intake | 1598 | Average weekly spirits intake |
| 1588 | Average weekly beer plus cider intake | 1578 | Average weekly champagne plus white wine intake | 1568 | Average weekly red wine intake |
| 1558 | Alcohol intake frequency | 399 | Number of incorrect matches in round | 94 | Diastolic blood pressure, manual reading |
| 93 | Systolic blood pressure, manual reading | 54 | UK Biobank assessment centre | 53 | Date of attending assessment centre |
| 48 | Waist circumference | 31 | Sex | | |

**Table S3: Cardiometabolic medication**

| Antihypertensive | | | | Lipid-lowering | Antidiabetic |
|---|---|---|---|---|---|
| hygroton | vasetic | acezide | capozide | clofibrate | glucobay |
| metoprolol | tenoret | tenoretic | sotalol | ciprofibrate | tolbutamide |
| timolol | beta-adalat | tenif | prestim | pravastatin | insulin |
| propranolol | acebutolol | atenolol | methyldopa | crestor | repaglinide |
| lisinopril | carace | zestril | quinapril | fenofibrate | nateglinide |
| accuretic | quinalapril | captopril | acepril | lescol | glipizide |
| capoten | innovace | innozide | enalapril | lipitor | glucophage |
| coversyl | ramipril | staril | cilazapril | ezetimibe | metformin |
| trandolapril | gopten | nifedipine | adalat | simvastatin | pioglitazone |
| coracten | tildiem | angiozem | adizem-60 | zocor | rosiglitazone |
| dilzem | cardene | isradipine | prescal | atorvastatin | glibenese |
| istin | lacidipine | motens | zestoretic | ezetrol | gliclazide |
| bisoprolol | hydroflumethiazide | methyclothiazide | polythiazide | lipostat | glimepiride |
| diurexan | aprinox | chlorothiazide | cyclopenthiazide | fluvastatin | actos |
| hydrochlorothiazide | triamterene | dyazide | dytide | rosuvastatin | acarbose |
| kalspare | moduret | moduretic | amiloride | | minodiab |
| nimodipine | amlodipine | diltiazem | nicardipine | | diamicron |
| oxprenolol | adalate | accupro | verapamil | | amaryl |
| fosinopril | perindopril | antihypertensive | felodipine | | starlix |
| carvedilol | slozem | beta-blocker | losartan | | bezafibrate |
| cozaar | angitil | metazem | co-triamterzide | | |
| co-amilozide | co-flumactone | co-tenidone | adipine | | |
| kentiazem | moexipril | perdix | adizem-xl | | |
| amilamont | tensipine | plendil | nisoldipine | | |
| syscor | valsartan | diovan | natrilix | | |
| slofedipine | verapress | cozaar-comp | hypapril | | |
| viazem | irbesartan | aprovel | lercanidipine | | |
| zanidip | tarka | candesartan | amias | | |
| nifedipress | imidapril | tanatril | nebivolol | | |
| triapin | telmisartan | tensopril | zemtard | | |
| cardicor | eprosartan | teveten | micardis | | |
| coaprovel | zemret | bi-carzem | horizem | | |
| micardisplus | felotens | tritace | felendil | | |
| vascalpha | olmesartan | olmetec | bendroflumethiazide | | |
| nadolol | cardioplen | lopace | amlostin | | |
| co-diovan | eplerenone | inspra | | | |

*Notes:* The table lists the unique generic and brand names of drugs registered in nurse-led interviews (data field 20003). The drugs are listed on their own, even if they were registered as part of combination drugs.

**Table S4: Cardiometabolic and cognitive outliers**

| A: Cardiometabolic risk factors | | B: Cognitive tests | |
|---|---|---|---|
| **Variables** | **Outliers** | **Variables** | **Outliers** |
| Body mass index | 361 | Numeric memory | 108 |
| Waist circumference | 143 | Fluid intelligence | 53 |
| Systolic blood pressure | 192 | Trail making test B | 203 |
| Diastolic blood pressure | 139 | Matrix test | 31 |
| Pulse pressure | 297 | Symbol digit substitution | 130 |
| C-reactive protein | 546 | Tower rearranging | 33 |
| Glycated hemoglobin | 473 | Paired associate learning | 0 |
| High-density lipoprotein cholesterol | 241 | Pairs matching | 18 |
| Low-density lipoprotein cholesterol | 129 | | |
| Total cholesterol | 144 | | |
| Triglycerides | 269 | | |

### Table S5: Descriptive statistics for male and female participants in the total sample

| Variables | Male (n=15,464) | Female (n=17,164) | Test statistic | P-value |
|---|---|---|---|---|
| **Demographic variables** | | | | |
| Age at baseline, Years[1] | 55.56±7.6 | 54.24±7.38 | 15.94 | $5.4x10^{-57}$ |
| Age at imaging Years[1] | 64.9±7.78 | 63.6±7.58 | 15.24 | $3x10^{-52}$ |
| Asian[2] | 240 (1.6%) | 182 (1.1%) | 15.02 | 0.00011 |
| Black[2] | 77 (0.5%) | 105 (0.6%) | 1.70 | 0.19 |
| Multiethnic[2] | 47 (0.3%) | 107 (0.6%) | 17.00 | $3.7x10^{-5}$ |
| Other ethnic background[2] | 68 (0.4%) | 97 (0.6%) | 2.30 | 0.13 |
| White[2] | 14,976 (96.8%) | 16,630 (96.9%) | 0.04 | 0.84 |
| Higher education[2] | 7,513 (48.6%) | 7,649 (44.6%) | 52.68 | $3.9x10^{-13}$ |
| Intermediate education[2] | 4,489 (29%) | 6,178 (36%) | 179.04 | $7.9x10^{-41}$ |
| **Cardiometabolic risk factors** | | | | |
| Body mass index, $kg/m^2$ [1] | 27.08±3.64 | 26.03±4.52 | 23.17 | $8.1x10^{-118}$ |
| Waist circumference, cm[1] | 94.52±9.99 | 81.68±11.13 | 109.90 | 0 |
| Systolic blood pressure, mmHg[1] | 138.81±16.41 | 131.29±17.91 | 39.55 | 0 |
| Diastolic blood pressure, mmHg[1] | 83.59±9.64 | 79.46±9.69 | 38.62 | $4.3x10^{-302}$ |
| Pulse pressure, mmHg[1] | 55.22±11.54 | 51.84±12.72 | 25.15 | $2.7x10^{-138}$ |
| C-reactive protein, mg/dL[1] | 1.94±3.34 | 2.14±3.76 | -5.05 | $4.4x10^{-7}$ |
| Glycated hemoglobin, mmol/mol[1] | 35.2±5.5 | 34.71±4.56 | 8.75 | $2.2x10^{-18}$ |
| High-density lipoprotein cholesterol, mmol/L[1] | 1.31±0.3 | 1.64±0.37 | -89.20 | 0 |
| Low-density lipoprotein cholesterol, mmol/L[1] | 3.57±0.82 | 3.59±0.82 | -2.08 | 0.038 |
| Total cholesterol, mmol/L[1] | 5.6±1.07 | 5.85±1.07 | -20.96 | $7x10^{-97}$ |
| Triglycerides, mmol/L[1] | 1.89±1.05 | 1.41±0.76 | 46.83 | 0 |
| Cardiometabolic diagnoses | | | | |
| Hypertension[2] | 8,275 (53.5%) | 6,190 (36.1%) | 1002.74 | $4.6x10^{-220}$ |
| Diabetes[2] | 484 (3.1%) | 259 (1.5%) | 95.31 | $1.6x10^{-22}$ |
| Dyslipidemia[2] | 9,433 (61.0%) | 7,932 (46.2%) | 713.84 | $2.9x10^{-157}$ |
| **Liver fat and steatotic liver diseases** | | | | |
| Liver fat, %[1] | 4.70±4.18 | 3.86±3.97 | 18.45 | $1.3x10^{-75}$ |
| Nonalcoholic fatty liver disease | 3,136 (20.3%) | 2,663 (15.5%) | 126.02 | $3x10^{-29}$ |
| Metabolic dysfunction-associated fatty liver disease[2] | 4,145 (26.8%) | 2,915 (17%) | 462.16 | $1.6x10^{-102}$ |
| Metabolic dysfunction-associated steatotic liver disease[2] | 3,069 (19.8%) | 2,579 (15%) | 131.72 | $1.7x10^{-30}$ |
| White matter hyperintensities | | | | |
| White matter hyperintensities, mL[1] | 5,566.27±7,103.38 | 4,607.57±6,188.41 | 12.9 | $3.6x10^{-38}$ |
| **Behavioral risk factors** | | | | |
| Current smoker[2] | 590 (3.8%) | 473 (2.8%) | 28.64 | $8.7x10^{-8}$ |
| Former smoker[2] | 5,693 (36.8%) | 5,310 (30.9%) | 125.48 | $4x10^{-29}$ |
| Alcohol consumption, g[1] | 145.07±143.48 | 69.39±79.13 | 58.10 | 0 |

[1] Mean ± SD; Welch two sample T-test
[2] n (proportion); Pearson’s Chi-squared test with Yates’ continuity correction

#### Table S6: Descriptive statistics of the total sample and the subsample

| Variables | Total sample (n=32,628) | Subsample (n=23,467) | Test statistic | P-value |
|---|---|---|---|---|
| **Demographic variables** | | | | |
| Age at baseline, Years[1] | 54.87±7.51 | 54.49±7.48 | 5.91 | $3.5x10^{-9}$ |
| Age at imaging Years[1] | 64.21±7.7 | 64.59±7.61 | -5.77 | $8.1x10^{-9}$ |
| Male[2] | 15,464 (47.4%) | 11,124 (47.4%) | 0.00 | 0.99 |
| Female[2] | 17,164 (53.6%) | 12,343 (53.6%) | | |
| Asian[2] | 422 (1.3%) | 269 (1.1%) | 2.31 | 0.13 |
| Black[2] | 182 (0.6%) | 129 (0.5%) | 0.00 | 0.94 |
| Multiethnic[2] | 154 (0.5%) | 111 (0.5%) | 0.00 | 1 |
| Other ethnic background[2] | 165 (0.5%) | 118 (0.5%) | 0.00 | 1 |
| White[2] | 31,606 (96.9%) | 22,776 (97.1%) | 1.56 | 0.21 |
| Higher education[2] | 15,162 (46.5%) | 11,312 (48.2%) | 16.41 | $5.1x10^{-5}$ |
| Intermediate education[2] | 10,667 (32.7%) | 7,533 (32.1%) | 2.16 | 0.14 |
| **Cardiometabolic risk factors** | | | | |
| Body mass index, $kg/m^2$ [1] | 26.53±4.16 | 26.45±4.13 | 1.99 | 0.047 |
| Waist circumference, cm[1] | 87.76±12.39 | 87.58±12.32 | 1.70 | 0.09 |
| Systolic blood pressure, mmHg[1] | 134.86±17.62 | 134.47±17.52 | 2.57 | 0.01 |
| Diastolic blood pressure, mmHg[1] | 81.42±9.88 | 81.37±9.91 | 0.57 | 0.57 |
| Pulse pressure, mmHg[1] | 53.44±12.29 | 53.1±12.13 | 3.24 | 0.0012 |
| C-reactive protein, mg/dL[1] | 2.04±3.57 | 1.99±3.49 | 1.73 | 0.084 |
| Glycated hemoglobin, mmol/mol[1] | 34.95±5.03 | 34.87±5.02 | 1.72 | 0.086 |
| High-density lipoprotein cholesterol, mmol/L[1] | 1.48±0.38 | 1.48±0.38 | -1.08 | 0.28 |
| Low-density lipoprotein cholesterol, mmol/L[1] | 3.58±0.82 | 3.58±0.82 | 0.61 | 0.54 |
| Total cholesterol, mmol/L[1] | 5.73±1.07 | 5.72±1.07 | 0.48 | 0.63 |
| Triglycerides, mmol/L[1] | 1.63±0.94 | 1.62±0.94 | 1.67 | 0.096 |
| Cardiometabolic diagnoses | | | | |
| Hypertension[2] | 14,465 (44.3%) | 10,214 (43.5%) | 3.59 | 0.058 |
| Diabetes[2] | 743 (2.3%) | 519 (2.2%) | 0.24 | 0.63 |
| Dyslipidemia[2] | 17,365 (53.2%) | 12,286 (52.4%) | 4.08 | 0.043 |
| **Liver fat and steatotic liver diseases** | | | | |
| Liver fat, %[1] | 4.26±4.09 | 4.25±3.96 | 0.39 | 0.7 |
| Nonalcoholic fatty liver disease | 5,799 (17.8%) | 4,124 (17.6%) | 0.36 | 0.55 |
| Metabolic dysfunction-associated fatty liver disease[2] | 7,060 (21.6%) | 4,974 (21.2%) | 1.56 | 0.21 |
| Metabolic dysfunction-associated steatotic liver disease[2] | 5,648 (17.3%) | 4,002 (17.1%) | 0.61 | 0.43 |
| White matter hyperintensities | | | | |
| White matter hyperintensities, mL[1] | 5,061.94±6,654.94 | 5,288.49±6,822 | -3.92 | $8.9x10^{-5}$ |
| **Behavioral risk factors** | | | | |
| Current smoker[2] | 1,063 (3.3%) | 705 (3%) | 2.80 | 0.094 |
| Former smoker[2] | 11,003 (33.7%) | 7,832 (33.4%) | 0.73 | 0.39 |
| Alcohol consumption, g[1] | 105.26±120.33 | 104.53±118.64 | 0.71 | 0.48 |

[1] Mean ± SD; Welch two sample T-test
[2] n (proportion); Pearson’s Chi-squared test with Yates’ continuity correction

### Table S7: Liver fat regression analyses

| Predictor | Partial correlation coefficient | 95% confidence interval | T-statistic | P-value |
|---|---|---|---|---|
| **Cardiometabolic risk factors**[1] | | | | |
| Body mass index | 0.40 | 0.39, 0.41 | 78.55 | 0 |
| Waist circumference | 0.41 | 0.4, 0.42 | 80.37 | 0 |
| Systolic blood pressure | 0.13 | 0.12, 0.14 | 23.53 | $1.9x10^{-121}$ |
| Diastolic blood pressure | 0.18 | 0.17, 0.19 | 33.77 | $8.9x10^{-246}$ |
| Pulse pressure | 0.03 | 0.02, 0.04 | 5.67 | $1.4x10^{-8}$ |
| C-reactive protein | 0.26 | 0.25, 0.27 | 47.81 | 0 |
| Glycated hemoglobin | 0.15 | 0.14, 0.16 | 27.76 | $1.1x10^{-167}$ |
| High-density lipoprotein cholesterol | -0.27 | -0.28, -0.26 | -50.46 | 0 |
| Low-density lipoprotein cholesterol | 0.08 | 0.07, 0.09 | 14.68 | $1.3x10^{-48}$ |
| Total cholesterol | 0.04 | 0.03, 0.05 | 6.97 | $3.3x10^{-12}$ |
| Triglycerides | 0.32 | 0.31, 0.33 | 61.65 | 0 |
| **Cardiometabolic principal components**[1] | | | | |
| Principal component 1 | 0.36 | 0.35, 0.37 | 69.23 | 0 |
| Principal component 2 | 0.13 | 0.12, 0.14 | 23.60 | $4.2x10^{-122}$ |
| Principal component 3 | -0.23 | -0.24, -0.22 | -42.08 | 0 |

*Notes:* Associations derived from multiple linear regression analyses between predictors (each predictor analyzed in a separate regression model) and the outcome variable liver fat.
[1]Adjusted for age, $age^2$, sex, age-by-sex, $age^2$-by-sex, assessment center, smoking status, and alcohol consumption.

**Table S8: Sex-by-cardiometabolic interaction effects on liver fat**

| Sex-by-predictor | Partial correlation coefficient | 95% confidence interval | T-statistic | P-value |
|---|---|---|---|---|
| **Cardiometabolic risk factors[1]** | | | | |
| Body mass index | 0.028 | 0.017, 0.039 | 5.049 | 4.5x10-07 |
| Waist circumference | -0.008 | -0.019, 0.003 | -1.459 | 0.14 |
| Systolic blood pressure | -0.018 | -0.028, -0.007 | -3.179 | 0.0015 |
| Diastolic blood pressure | -0.024 | -0.034, -0.013 | -4.249 | 2.2x10-05 |
| Pulse pressure | -0.014 | -0.025, -0.004 | -2.595 | 0.0095 |
| C-reactive protein | -0.047 | -0.058, -0.036 | -8.536 | 1.5x10-17 |
| Glycated hemoglobin | -0.017 | -0.028, -0.006 | -3.073 | 0.0021 |
| High-density lipoprotein cholesterol | -0.012 | -0.023, -0.001 | -2.165 | 0.03 |
| Low-density lipoprotein cholesterol | -0.031 | -0.042, -0.02 | -5.600 | 2.2x10-08 |
| Total cholesterol | -0.011 | -0.022, 0 | -1.962 | 0.05 |
| Triglycerides | -0.070 | -0.081, -0.059 | -12.696 | 7.7x10-37 |
| **Cardiometabolic principal components[1]** | | | | |
| Principal component 1 | -0.011 | -0.022, 0 | -1.982 | 0.048 |
| Principal component 2 | -0.015 | -0.026, -0.005 | -2.797 | 0.0052 |
| Principal component 3 | 0.012 | 0.001, 0.022 | 2.086 | 0.037 |

*Notes:* Interaction effects derived from multiple linear regression analyses between predictors (each predictor analyzed in a separate regression model) and sex (predictor-by-sex) on the outcome variable liver fat.
[1]Adjusted for age, age$^2$, sex, age-by-sex, age$^2$-by-sex, assessment center, smoking status, and alcohol consumption.

**Table S9: White matter hyperintensities regression analyses**

| Predictor | Partial correlation coefficient | 95% confidence interval | T-statistic | P-value |
|---|---|---|---|---|
| **Cardiometabolic risk factors[1]** | | | | |
| Body mass index | 0.121 | 0.11, 0.132 | 22.022 | $1.1x10^{-106}$ |
| Waist circumference | 0.122 | 0.111, 0.133 | 22.186 | $3x10^{-108}$ |
| Systolic blood pressure | 0.112 | 0.101, 0.123 | 20.329 | $2.6x10^{-91}$ |
| Diastolic blood pressure | 0.129 | 0.119, 0.14 | 23.545 | $1.5x10^{-121}$ |
| Pulse pressure | 0.053 | 0.042, 0.063 | 9.513 | $2x10^{-21}$ |
| C-reactive protein | 0.052 | 0.041, 0.063 | 9.400 | $5.8x10^{-21}$ |
| Glycated hemoglobin | 0.054 | 0.043, 0.064 | 9.691 | $3.5x10^{-22}$ |
| High-density lipoprotein cholesterol | -0.047 | -0.058, -0.036 | -8.543 | $1.4x10^{-17}$ |
| Low-density lipoprotein cholesterol | -0.002 | -0.012, 0.009 | -0.292 | 0.77 |
| Total cholesterol | -0.009 | -0.02, 0.002 | -1.578 | 0.11 |
| Triglycerides | 0.041 | 0.03, 0.052 | 7.391 | $1.5x10^{-13}$ |
| **Cardiometabolic principal components[1]** | | | | |
| Principal component 1 | 0.143 | 0.132, 0.153 | 26.070 | $2.7x10^{-148}$ |
| Principal component 2 | 0.046 | 0.035, 0.057 | 8.347 | $7.3x10^{-17}$ |
| Principal component 3 | 0.006 | -0.005, 0.017 | 1.089 | 0.28 |
| **Liver fat and steatotic liver diseases[1]** | | | | |
| Liver fat | 0.094 | 0.083, 0.104 | 16.961 | $3x10^{-64}$ |
| Nonalcoholic fatty liver disease | 0.066 | 0.055, 0.077 | 11.931 | $9.6x10^{-33}$ |
| Metabolic dysfunction-associated fatty liver disease | 0.078 | 0.068, 0.089 | 14.200 | $1.2x10^{-45}$ |
| Metabolic dysfunction-associated steatotic liver disease | 0.067 | 0.056, 0.078 | 12.174 | $5x10^{-34}$ |

*Notes:* Associations derived from multiple linear regression analyses between predictors (each predictor analyzed in a separate regression model) and the outcome variable white matter hyperintensities.

[1]Adjusted for age, $age^2$, sex, age-by-sex, $age^2$-by-sex, assessment center, smoking status, alcohol consumption, and intracranial volume.

### Table S10: Interaction effects with sex on white matter hyperintensities

| Sex-by-predictor | Partial correlation coefficient | 95% confidence interval | T-statistic | P-value |
|---|---|---|---|---|
| **Cardiometabolic risk factors**[1] | | | | |
| Body mass index | 0.047 | 0.036, 0.057 | 8.438 | $3.3x10^{-17}$ |
| Waist circumference | 0.036 | 0.026, 0.047 | 6.594 | $4.4x10^{-11}$ |
| Systolic blood pressure | -0.008 | -0.019, 0.003 | -1.468 | 0.14 |
| Diastolic blood pressure | -0.011 | -0.022, 0 | -1.994 | 0.046 |
| Pulse pressure | -0.008 | -0.019, 0.003 | -1.507 | 0.13 |
| C-reactive protein | 0.008 | -0.003, 0.019 | 1.484 | 0.14 |
| Glycated hemoglobin | 0.000 | -0.011, 0.011 | 0.004 | 1 |
| High-density lipoprotein cholesterol | -0.021 | -0.032, -0.01 | -3.809 | 0.00014 |
| Low-density lipoprotein cholesterol | -0.014 | -0.025, -0.003 | -2.561 | 0.01 |
| Total cholesterol | -0.014 | -0.024, -0.003 | -2.464 | 0.014 |
| Triglycerides | -0.004 | -0.015, 0.007 | -0.737 | 0.46 |
| **Cardiometabolic principal components**[1] | | | | |
| Principal component 1 | 0.016 | 0.005, 0.027 | 2.836 | 0.0046 |
| Principal component 2 | 0.022 | 0.011, 0.033 | 3.970 | $7.2x10^{-05}$ |
| Principal component 3 | -0.020 | -0.031, -0.01 | -3.686 | 0.00023 |
| **Liver fat and steatotic liver diseases**[1] | | | | |
| Liver fat | 0.014 | 0.003, 0.025 | 2.552 | 0.011 |
| Nonalcoholic fatty liver disease | 0.006 | -0.005, 0.017 | 1.090 | 0.28 |
| Metabolic dysfunction-associated fatty liver disease | 0.011 | 0, 0.022 | 1.948 | 0.051 |
| Metabolic dysfunction-associated steatotic liver disease | 0.007 | -0.004, 0.018 | 1.230 | 0.22 |

*Notes:* Interaction effects derived from multiple linear regression analyses between predictors (each predictor analyzed in a separate regression model) and sex (predictor-by-sex) on the outcome variable white matter hyperintensities.
[1]Adjusted for age, $age^2$, sex, age-by-sex, $age^2$-by-sex, assessment center, smoking status, alcohol consumption, and intracranial volume.

**Table S11: The cognitive principal component 1 regression analyses**

| Predictor | Partial correlation coefficient | 95% confidence interval | T-statistic | P-value |
|---|---|---|---|---|
| **Cardiometabolic risk factors[1]** | | | | |
| Body mass index | -0.032 | -0.043, -0.021 | -4.941 | $7.8 \times 10^{-7}$ |
| Waist circumference | -0.021 | -0.031, -0.01 | -3.151 | 0.0016 |
| Systolic blood pressure | -0.027 | -0.038, -0.016 | -4.113 | $3.9 \times 10^{-5}$ |
| Diastolic blood pressure | -0.028 | -0.039, -0.017 | -4.255 | $2.1 \times 10^{-5}$ |
| Pulse pressure | -0.015 | -0.026, -0.004 | -2.344 | 0.019 |
| C-reactive protein | -0.016 | -0.027, -0.005 | -2.481 | 0.013 |
| Glycated hemoglobin | -0.037 | -0.048, -0.026 | -5.702 | $1.2 \times 10^{-8}$ |
| High-density lipoprotein cholesterol | 0.012 | 0.001, 0.023 | 1.891 | 0.059 |
| Low-density lipoprotein cholesterol | 0.003 | -0.007, 0.014 | 0.528 | 0.6 |
| Total cholesterol | 0.004 | -0.007, 0.015 | 0.563 | 0.57 |
| Triglycerides | -0.021 | -0.031, -0.01 | -3.141 | 0.0017 |
| **Cardiometabolic principal components[1]** | | | | |
| Principal component 1 | -0.035 | -0.046, -0.024 | -5.330 | $9.9 \times 10^{-8}$ |
| Principal component 2 | -0.013 | -0.023, -0.002 | -1.920 | 0.055 |
| Principal component 3 | -0.003 | -0.014, 0.008 | -0.443 | 0.66 |
| **Liver fat and steatotic liver disease[1]** | | | | |
| Liver fat | -0.034 | -0.045, -0.023 | -5.253 | $1.5 \times 10^{-7}$ |
| Nonalcoholic fatty liver disease | -0.031 | -0.042, -0.02 | -4.759 | $2 \times 10^{-6}$ |
| Metabolic dysfunction-associated fatty liver disease | -0.032 | -0.043, -0.021 | -4.853 | $1.2 \times 10^{-6}$ |
| Metabolic dysfunction-associated steatotic liver disease | -0.030 | -0.041, -0.019 | -4.638 | $3.5 \times 10^{-6}$ |
| White matter hyperintensities[2] | | | | |
| White matter hyperintensities | -0.071 | -0.082, -0.06 | -10.912 | $1.2 \times 10^{-27}$ |

*Notes:* Associations derived from multiple linear regression analyses between predictors (each predictor analyzed in a separate regression model) and the outcome variable the cognitive principal component 1.

[1]Adjusted for age, $age^2$, sex, age-by-sex, $age^2$-by-sex, assessment center, smoking status, alcohol consumption, and education.

[2]Adjusted for age, $age^2$, sex, age-by-sex, $age^2$-by-sex, assessment center, smoking status, alcohol consumption, education, and intracranial volume.

**Table S12: Interaction effects with sex on cognitive principal component 1**

| Sex-by-predictor | Partial correlation coefficient | 95% confidence interval | T-statistic | P-value |
|---|---|---|---|---|
| **Cardiometabolic risk factors[1]** | | | | |
| Body mass index | -0.022 | -0.032, -0.011 | -3.311 | 0.00093 |
| Waist circumference | -0.009 | -0.02, 0.002 | -1.426 | 0.15 |
| Systolic blood pressure | -0.003 | -0.014, 0.007 | -0.527 | 0.6 |
| Diastolic blood pressure | -0.004 | -0.015, 0.007 | -0.591 | 0.55 |
| Pulse pressure | 0.000 | -0.011, 0.011 | -0.048 | 0.96 |
| C-reactive protein | -0.001 | -0.011, 0.01 | -0.099 | 0.92 |
| Glycated hemoglobin | -0.007 | -0.018, 0.004 | -1.065 | 0.29 |
| High-density lipoprotein cholesterol | 0.003 | -0.008, 0.014 | 0.449 | 0.65 |
| Low-density lipoprotein cholesterol | 0.010 | -0.001, 0.02 | 1.461 | 0.14 |
| Total cholesterol | 0.009 | -0.002, 0.019 | 1.319 | 0.19 |
| Triglycerides | 0.005 | -0.006, 0.016 | 0.782 | 0.43 |
| **Cardiometabolic principal components[1]** | | | | |
| Principal component 1 | -0.006 | -0.017, 0.004 | -0.994 | 0.32 |
| Principal component 2 | -0.010 | -0.021, 0.001 | -1.481 | 0.14 |
| Principal component 3 | 0.001 | -0.01, 0.012 | 0.173 | 0.86 |
| **Liver fat and steatotic liver disease[1]** | | | | |
| Liver fat | 0.007 | -0.004, 0.018 | 1.104 | 0.27 |
| Nonalcoholic fatty liver disease | 0.004 | -0.007, 0.014 | 0.550 | 0.58 |
| Metabolic dysfunction-associated fatty liver disease | -0.003 | -0.014, 0.007 | -0.533 | 0.59 |
| Metabolic dysfunction-associated steatotic liver disease | 0.002 | -0.009, 0.013 | 0.290 | 0.77 |
| **White matter hyperintensities[2]** | | | | |
| White matter hyperintensities | -0.008 | -0.019, 0.003 | -1.193 | 0.23 |

*Notes:* Interaction effects derived from multiple linear regression analyses between predictors (each predictor analyzed in a separate regression model) and sex (predictor-by-sex) on the outcome variable the cognitive principal component 1.

[1]Adjusted for age, age$^2$, sex, age-by-sex, age$^2$-by-sex, assessment center, smoking status, alcohol consumption, and education.

[2]Adjusted for age, age$^2$, sex, age-by-sex, age$^2$-by-sex, assessment center, smoking status, alcohol consumption, education, and intracranial volume.

**Table S13: Regression with individual cognitive tests**

| | Partial correlation coefficient | 95% confidence interval | T-statistic | P-value | Partial correlation coefficient | 95% confidence interval | T-statistic | P-value |
|---|---|---|---|---|---|---|---|---|
| **Numeric memory** | | | | | **Fluid intelligence** | | | |
| **Cardiometabolic risk factors[1]** | | | | | | | | |
| Body mass index | -0.053 | -0.065, -0.040 | -8.126 | $4.7\times10^{-16}$ | -0.003 | -0.014, 0.009 | -0.458 | 0.65 |
| Waist circumference | -0.042 | -0.054, -0.029 | -6.445 | $1.2\times10^{-10}$ | 0.011 | -0.001, 0.022 | 1.865 | 0.062 |
| Systolic blood pressure | -0.036 | -0.048, -0.023 | -5.505 | $3.7\times10^{-8}$ | -0.011 | -0.022, 0 | -1.946 | 0.052 |
| Diastolic blood pressure | -0.038 | -0.051, -0.025 | -5.834 | $5.5\times10^{-9}$ | -0.021 | -0.032, -0.01 | -3.615 | $3\times10^{-4}$ |
| Pulse pressure | -0.020 | -0.032, -0.007 | -3.021 | 0.0025 | 0.002 | -0.01, 0.013 | 0.265 | 0.79 |
| C-reactive protein | -0.021 | -0.034, -0.009 | -3.313 | 0.0009 | -0.006 | -0.018, 0.005 | -1.096 | 0.27 |
| Glycated hemoglobin | -0.023 | -0.036, -0.011 | -3.612 | $3\times10^{-4}$ | -0.009 | -0.021, 0.002 | -1.630 | 0.1 |
| High-density lipoprotein cholesterol | 0.027 | 0.014, 0.04 | 4.137 | $3.5\times10^{-5}$ | 0.005 | -0.006, 0.016 | 0.896 | 0.37 |
| Low-density lipoprotein cholesterol | -0.006 | -0.019, 0.007 | -0.941 | 0.35 | 0.001 | -0.01, 0.013 | 0.231 | 0.82 |
| Total cholesterol | -0.002 | -0.015, 0.011 | -0.311 | 0.76 | 0.002 | -0.009, 0.014 | 0.429 | 0.67 |
| Triglycerides | -0.035 | -0.048, -0.023 | -5.429 | $5.7\times10^{-8}$ | 0.003 | -0.008, 0.015 | 0.608 | 0.54 |
| **Cardiometabolic principal components[1]** | | | | | | | | |
| Principal component 1 | -0.054 | -0.067, -0.041 | -8.355 | $6.9\times10^{-17}$ | -0.008 | -0.02, 0.003 | -1.470 | 0.14 |
| Principal component 2 | -0.014 | -0.026, -0.001 | -2.084 | 0.037 | -0.003 | -0.014, 0.009 | -0.469 | 0.64 |
| Principal component 3 | 0.009 | -0.004, 0.022 | 1.397 | 0.16 | -0.008 | -0.019, 0.003 | -1.388 | 0.17 |
| **Liver fat and steatotic liver disease[1]** | | | | | | | | |
| Liver fat | -0.046 | -0.058, -0.033 | -7.038 | $2\times10^{-12}$ | -0.022 | -0.033, -0.01 | -3.743 | 0.0002 |
| Nonalcoholic fatty liver disease | -0.036 | -0.048, -0.023 | -5.478 | $4.3\times10^{-8}$ | -0.010 | -0.021, 0.001 | -1.742 | 0.081 |
| Metabolic dysfunction-associated fatty liver disease | -0.039 | -0.052, -0.027 | -6.085 | $1.2\times10^{-9}$ | -0.013 | -0.025, -0.002 | -2.304 | 0.021 |
| Metabolic dysfunction-associated steatotic liver disease | -0.035 | -0.047, -0.022 | -5.362 | $8.3\times10^{-8}$ | -0.009 | -0.02, 0.002 | -1.559 | 0.12 |
| **White matter hyperintensities[2]** | | | | | | | | |
| White matter hyperintensities | -0.037 | -0.05, -0.025 | -5.748 | $9.1\times10^{-9}$ | -0.044 | -0.055, -0.033 | -7.682 | $1.6\times10^{-14}$ |
| **Trail making test B** | | | | | **Matrix test** | | | |
| **Cardiometabolic risk factors[1]** | | | | | | | | |
| Body mass index | 0.006 | -0.008, 0.019 | 0.785 | 0.43 | -0.019 | -0.033, -0.005 | -2.723 | 0.0065 |
| Waist circumference | -0.004 | -0.018, 0.01 | -0.555 | 0.58 | -0.009 | -0.023, 0.005 | -1.311 | 0.19 |
| Systolic blood pressure | 0.005 | -0.009, 0.018 | 0.654 | 0.51 | -0.011 | -0.025, 0.002 | -1.619 | 0.11 |
| Diastolic blood pressure | 0.008 | -0.006, 0.022 | 1.167 | 0.24 | -0.012 | -0.025, 0.002 | -1.680 | 0.093 |
| Pulse pressure | 0.000 | -0.014, 0.013 | -0.057 | 0.95 | -0.006 | -0.02, 0.007 | -0.916 | 0.36 |
| C-reactive protein | 0.011 | -0.003, 0.025 | 1.597 | 0.11 | -0.006 | -0.019, 0.008 | -0.795 | 0.43 |
| Glycated hemoglobin | 0.026 | 0.013, 0.04 | 3.732 | 0.0002 | -0.015 | -0.029, -0.002 | -2.176 | 0.03 |
| High-density lipoprotein cholesterol | 0.003 | -0.011, 0.017 | 0.421 | 0.67 | 0.000 | -0.014, 0.013 | -0.062 | 0.95 |
| Low-density lipoprotein cholesterol | -0.005 | -0.019, 0.009 | -0.742 | 0.46 | 0.003 | -0.011, 0.016 | 0.378 | 0.71 |
| Total cholesterol | -0.003 | -0.017, 0.011 | -0.430 | 0.67 | 0.003 | -0.01, 0.017 | 0.471 | 0.64 |
| Triglycerides | -0.005 | -0.019, 0.009 | -0.732 | 0.46 | -0.002 | -0.016, 0.011 | -0.343 | 0.73 |
| **Cardiometabolic principal components[1]** | | | | | | | | |
| Principal component 1 | 0.005 | -0.009, 0.019 | 0.743 | 0.46 | -0.015 | -0.028, -0.001 | -2.084 | 0.037 |
| Principal component 2 | 0.003 | -0.011, 0.016 | 0.375 | 0.71 | -0.008 | -0.021, 0.006 | -1.110 | 0.27 |
| Principal component 3 | 0.006 | -0.008, 0.02 | 0.889 | 0.37 | -0.003 | -0.016, 0.011 | -0.389 | 0.7 |
| **Liver fat and steatotic liver disease[1]** | | | | | | | | |
| Liver fat | 0.004 | -0.01, 0.017 | 0.519 | 0.6 | -0.030 | -0.043, -0.016 | -4.240 | $2.2\times10^{-5}$ |
| Nonalcoholic fatty liver disease | 0.005 | -0.009, 0.018 | 0.641 | 0.52 | -0.029 | -0.043, -0.016 | -4.224 | $2.4\times10^{-5}$ |
| Metabolic dysfunction-associated fatty liver disease | 0.006 | -0.008, 0.019 | 0.795 | 0.43 | -0.023 | -0.037, -0.009 | -3.283 | 0.001 |
| Metabolic dysfunction-associated steatotic liver disease | 0.006 | -0.008, 0.02 | 0.847 | 0.4 | -0.029 | -0.043, -0.015 | -4.141 | $3.5\times10^{-5}$ |
| **White matter hyperintensities[2]** | | | | | | | | |
| White matter hyperintensities | 0.052 | 0.038, 0.066 | 7.399 | $1.4\times10^{-13}$ | -0.052 | -0.065, -0.038 | -7.411 | $1.3\times10^{-13}$ |
| **Symbol digit substitution** | | | | | **Tower rearranging** | | | |
| **Cardiometabolic risk factors[1]** | | | | | | | | |
| Body mass index | -0.040 | -0.053, -0.026 | -5.671 | $1.4\times10^{-8}$ | 0.030 | 0.016, 0.044 | 4.275 | $1.9\times10^{-5}$ |

| | Partial correlation coefficient | 95% confidence interval | T-statistic | P-value | Partial correlation coefficient | 95% confidence interval | T-statistic | P-value |
|---|---|---|---|---|---|---|---|---|
| Waist circumference | -0.039 | -0.052, -0.025 | -5.542 | $3x10^{-8}$ | 0.034 | 0.021, 0.048 | 4.925 | $8.5x10^{-7}$ |
| Systolic blood pressure | -0.010 | -0.023, 0.004 | -1.382 | 0.17 | -0.001 | -0.014, 0.013 | -0.078 | 0.94 |
| Diastolic blood pressure | -0.021 | -0.034, -0.007 | -2.991 | 0.0028 | 0.011 | -0.003, 0.025 | 1.569 | 0.12 |
| Pulse pressure | 0.004 | -0.01, 0.018 | 0.571 | 0.57 | -0.010 | -0.024, 0.003 | -1.480 | 0.14 |
| C-reactive protein | -0.022 | -0.035, -0.008 | -3.118 | 0.0018 | 0.021 | 0.008, 0.035 | 3.035 | 0.0024 |
| Glycated hemoglobin | -0.042 | -0.056, -0.028 | -6.027 | $1.7x10^{-9}$ | -0.019 | -0.033, -0.005 | -2.683 | 0.0073 |
| High-density lipoprotein cholesterol | 0.015 | 0.001, 0.029 | 2.163 | 0.031 | -0.015 | -0.028, -0.001 | -2.109 | 0.035 |
| Low-density lipoprotein cholesterol | 0.002 | -0.011, 0.016 | 0.319 | 0.75 | 0.005 | -0.009, 0.019 | 0.711 | 0.48 |
| Total cholesterol | 0.002 | -0.012, 0.015 | 0.258 | 0.8 | 0.001 | -0.013, 0.014 | 0.089 | 0.93 |
| Triglycerides | -0.026 | -0.04, -0.012 | -3.752 | 0.0002 | 0.009 | -0.005, 0.023 | 1.278 | 0.2 |
| **Cardiometabolic principal components[1]** | | | | | | | | |
| Principal component 1 | -0.031 | -0.044, -0.017 | -4.387 | $1.2x10^{-5}$ | 0.019 | 0.005, 0.033 | 2.739 | 0.0062 |
| Principal component 2 | -0.016 | -0.029, -0.002 | -2.224 | 0.026 | 0.012 | -0.002, 0.026 | 1.745 | 0.081 |
| Principal component 3 | 0.017 | 0.003, 0.031 | 2.439 | 0.015 | -0.023 | -0.037, -0.01 | -3.339 | 0.0008 |
| **Liver fat and steatotic liver disease[1]** | | | | | | | | |
| Liver fat | -0.030 | -0.044, -0.017 | -4.350 | $1.4x10^{-5}$ | 0.023 | 0.01, 0.037 | 3.326 | 0.0009 |
| Nonalcoholic fatty liver disease | -0.031 | -0.045, -0.017 | -4.463 | $8.1x10^{-6}$ | 0.014 | 0.001, 0.028 | 2.048 | 0.041 |
| Metabolic dysfunction-associated fatty liver disease | -0.033 | -0.047, -0.02 | -4.770 | $1.9x10^{-6}$ | 0.022 | 0.008, 0.036 | 3.167 | 0.0015 |
| Metabolic dysfunction-associated steatotic liver disease | -0.031 | -0.045, -0.017 | -4.449 | $8.7x10^{-6}$ | 0.016 | 0.002, 0.029 | 2.218 | 0.027 |
| **White matter hyperintensities[2]** | | | | | | | | |
| White matter hyperintensities | -0.061 | -0.074, -0.047 | -8.708 | $3.3x10^{-18}$ | -0.034 | -0.047, -0.02 | -4.808 | $1.5x10^{-6}$ |
| **Paired associate learning** | | | | | **Pairs matching** | | | |
| **Cardiometabolic risk factors[1]** | | | | | | | | |
| Body mass index | -0.056 | -0.069, -0.043 | -8.649 | $5.5x10^{-18}$ | -0.031 | -0.043, -0.02 | -5.523 | $3.4x10^{-8}$ |
| Waist circumference | -0.050 | -0.063, -0.038 | -7.770 | $8.2x10^{-15}$ | -0.033 | -0.044, -0.022 | -5.816 | $6.1x10^{-9}$ |
| Systolic blood pressure | -0.014 | -0.027, -0.002 | -2.228 | 0.026 | -0.006 | -0.018, 0.005 | -1.120 | 0.26 |
| Diastolic blood pressure | -0.019 | -0.032, -0.007 | -2.970 | 0.003 | -0.010 | -0.021, 0.001 | -1.698 | 0.089 |
| Pulse pressure | -0.005 | -0.017, 0.008 | -0.699 | 0.48 | -0.001 | -0.012, 0.01 | -0.177 | 0.86 |
| C-reactive protein | -0.025 | -0.037, -0.012 | -3.794 | 0.0002 | -0.013 | -0.024, -0.001 | -2.199 | 0.028 |
| Glycated hemoglobin | -0.026 | -0.039, -0.013 | -4.031 | $5.6x10^{-5}$ | 0.004 | -0.008, 0.015 | 0.637 | 0.52 |
| High-density lipoprotein cholesterol | 0.019 | 0.006, 0.032 | 2.902 | 0.0037 | 0.010 | -0.001, 0.021 | 1.708 | 0.088 |
| Low-density lipoprotein cholesterol | -0.005 | -0.018, 0.008 | -0.756 | 0.45 | -0.021 | -0.032, -0.01 | -3.654 | 0.00026 |
| Total cholesterol | -0.004 | -0.016, 0.009 | -0.538 | 0.59 | -0.017 | -0.029, -0.006 | -3.065 | 0.0022 |
| Triglycerides | -0.032 | -0.044, -0.019 | -4.879 | $1.1x10^{-6}$ | -0.014 | -0.025, -0.003 | -2.411 | 0.016 |
| **Cardiometabolic principal components[1]** | | | | | | | | |
| Principal component 1 | -0.043 | -0.056, -0.03 | -6.648 | $3x10^{-11}$ | -0.027 | -0.038, -0.016 | -4.686 | $2.8x10^{-6}$ |
| Principal component 2 | -0.015 | -0.028, -0.003 | -2.371 | 0.018 | 0.006 | -0.005, 0.017 | 1.055 | 0.29 |
| Principal component 3 | 0.026 | 0.013, 0.039 | 4.015 | $6x10^{-5}$ | 0.025 | 0.014, 0.036 | 4.391 | $1.1x10^{-5}$ |
| **Liver fat and steatotic liver disease[1]** | | | | | | | | |
| Liver fat | -0.042 | -0.055, -0.03 | -6.535 | $6.5x10^{-11}$ | -0.018 | -0.029, -0.007 | -3.112 | 0.0019 |
| Nonalcoholic fatty liver disease | -0.037 | -0.05, -0.025 | -5.744 | $9.4x10^{-9}$ | -0.016 | -0.027, -0.005 | -2.799 | 0.0051 |
| Metabolic dysfunction-associated fatty liver disease | -0.043 | -0.056, -0.031 | -6.658 | $2.8x10^{-11}$ | -0.020 | -0.031, -0.009 | -3.519 | 0.0004 |
| Metabolic dysfunction-associated steatotic liver disease | -0.039 | -0.051, -0.026 | -5.942 | $2.9x10^{-9}$ | -0.018 | -0.029, -0.007 | -3.193 | 0.0014 |
| **White matter hyperintensities[2]** | | | | | | | | |
| White matter hyperintensities | -0.047 | -0.059, -0.034 | -7.160 | $8.3x10^{-13}$ | 0.017 | 0.006, 0.029 | 3.055 | 0.0023 |

*Notes:* Associations derived from multiple linear regression analyses between predictors (each predictor analyzed in a separate regression model) and the outcomes (the individual cognitive tests).

[1]Adjusted for age, age$^2$, sex, age-by-sex, age$^2$-by-sex, assessment center, smoking status, alcohol consumption, and education.

[2]Adjusted for age, age$^2$, sex, age-by-sex, age$^2$-by-sex, assessment center, smoking status, alcohol consumption, education, and intracranial volume.

**Table S14: Interaction effects with sex on cognitive tests**

| | Partial correlation coefficient | 95% confidence interval | T-statistic | P-value | Partial correlation coefficient | 95% confidence interval | T-statistic | P-value |
|---|---|---|---|---|---|---|---|---|
| **Numeric memory** | | | | | **Fluid intelligence** | | | |
| **Cardiometabolic risk factors**[1] | | | | | | | | |
| Body mass index | -0.015 | -0.027, -0.002 | -2.268 | 0.023 | -0.018 | -0.029, -0.006 | -3.081 | 0.0021 |
| Waist circumference | -0.007 | -0.02, 0.005 | -1.152 | 0.25 | -0.011 | -0.022, 0 | -1.918 | 0.055 |
| Systolic blood pressure | -0.005 | -0.017, 0.008 | -0.733 | 0.46 | -0.005 | -0.016, 0.006 | -0.872 | 0.38 |
| Diastolic blood pressure | -0.007 | -0.02, 0.005 | -1.127 | 0.26 | -0.006 | -0.018, 0.005 | -1.121 | 0.26 |
| Pulse pressure | 0.001 | -0.011, 0.014 | 0.200 | 0.84 | -0.001 | -0.012, 0.011 | -0.118 | 0.91 |
| C-reactive protein | -0.007 | -0.02, 0.006 | -1.091 | 0.28 | -0.004 | -0.015, 0.007 | -0.717 | 0.47 |
| Glycated hemoglobin | -0.007 | -0.02, 0.005 | -1.134 | 0.26 | -0.014 | -0.025, -0.002 | -2.392 | 0.017 |
| High-density lipoprotein cholesterol | -0.001 | -0.013, 0.012 | -0.101 | 0.92 | -0.001 | -0.012, 0.01 | -0.159 | 0.87 |
| Low-density lipoprotein cholesterol | -0.008 | -0.02, 0.005 | -1.182 | 0.24 | 0.006 | -0.005, 0.017 | 1.017 | 0.31 |
| Total cholesterol | -0.010 | -0.023, 0.003 | -1.534 | 0.12 | 0.003 | -0.008, 0.015 | 0.590 | 0.56 |
| Triglycerides | 0.002 | -0.011, 0.015 | 0.321 | 0.75 | 0.000 | -0.012, 0.011 | -0.051 | 0.96 |
| **Cardiometabolic principal components**[1] | | | | | | | | |
| Principal component 1 | -0.009 | -0.022, 0.003 | -1.438 | 0.15 | -0.010 | -0.021, 0.002 | -1.664 | 0.096 |
| Principal component 2 | 0.008 | -0.004, 0.021 | 1.303 | 0.19 | -0.007 | -0.018, 0.004 | -1.250 | 0.21 |
| Principal component 3 | 0.007 | -0.006, 0.019 | 1.014 | 0.31 | 0.003 | -0.009, 0.014 | 0.454 | 0.65 |
| **Liver fat and steatotic liver disease**[1] | | | | | | | | |
| Liver fat | -0.002 | -0.015, 0.01 | -0.382 | 0.7 | 0.009 | -0.002, 0.02 | 1.561 | 0.12 |
| Nonalcoholic fatty liver disease | -0.006 | -0.018, 0.007 | -0.873 | 0.38 | 0.007 | -0.004, 0.018 | 1.179 | 0.24 |
| Metabolic dysfunction-associated fatty liver disease | -0.008 | -0.021, 0.005 | -1.261 | 0.21 | 0.000 | -0.011, 0.012 | 0.048 | 0.96 |
| Metabolic dysfunction-associated steatotic liver disease | -0.007 | -0.02, 0.006 | -1.071 | 0.28 | 0.005 | -0.006, 0.017 | 0.932 | 0.35 |
| **White matter hyperintensities**[2] | | | | | | | | |
| White matter hyperintensities | 0.002 | -0.01, 0.015 | 0.353 | 0.72 | -0.011 | -0.023, 0 | -1.978 | 0.048 |
| **Trail making test B** | | | | | **Matrix test** | | | |
| **Cardiometabolic risk factors**[1] | | | | | | | | |
| Body mass index | 0.011 | -0.003, 0.024 | 1.498 | 0.13 | -0.014 | -0.027, 0 | -1.979 | 0.048 |
| Waist circumference | 0.007 | -0.006, 0.021 | 1.054 | 0.29 | -0.008 | -0.022, 0.005 | -1.209 | 0.23 |
| Systolic blood pressure | -0.001 | -0.015, 0.013 | -0.112 | 0.91 | 0.002 | -0.012, 0.015 | 0.230 | 0.82 |
| Diastolic blood pressure | -0.002 | -0.016, 0.012 | -0.311 | 0.76 | 0.001 | -0.013, 0.014 | 0.078 | 0.94 |
| Pulse pressure | 0.000 | -0.013, 0.014 | 0.047 | 0.96 | 0.002 | -0.011, 0.016 | 0.358 | 0.72 |
| C-reactive protein | 0.008 | -0.006, 0.022 | 1.102 | 0.27 | 0.007 | -0.007, 0.02 | 0.936 | 0.35 |
| Glycated hemoglobin | 0.006 | -0.008, 0.02 | 0.848 | 0.4 | -0.008 | -0.022, 0.006 | -1.128 | 0.26 |
| High-density lipoprotein cholesterol | 0.000 | -0.014, 0.014 | 0.023 | 0.98 | 0.006 | -0.007, 0.02 | 0.921 | 0.36 |
| Low-density lipoprotein cholesterol | 0.000 | -0.014, 0.014 | 0.008 | 0.99 | 0.006 | -0.008, 0.02 | 0.850 | 0.4 |
| Total cholesterol | 0.000 | -0.014, 0.014 | -0.047 | 0.96 | 0.008 | -0.005, 0.022 | 1.202 | 0.23 |
| Triglycerides | 0.001 | -0.013, 0.015 | 0.131 | 0.9 | 0.004 | -0.01, 0.018 | 0.595 | 0.55 |
| **Cardiometabolic principal components**[1] | | | | | | | | |
| Principal component 1 | 0.004 | -0.01, 0.018 | 0.534 | 0.59 | -0.001 | -0.015, 0.012 | -0.198 | 0.84 |
| Principal component 2 | 0.001 | -0.013, 0.015 | 0.146 | 0.88 | -0.009 | -0.022, 0.005 | -1.224 | 0.22 |
| Principal component 3 | -0.002 | -0.016, 0.012 | -0.305 | 0.76 | 0.005 | -0.008, 0.019 | 0.767 | 0.44 |
| **Liver fat and steatotic liver disease**[1] | | | | | | | | |
| Liver fat | 0.001 | -0.013, 0.015 | 0.097 | 0.92 | 0.004 | -0.01, 0.018 | 0.562 | 0.57 |
| Nonalcoholic fatty liver disease | 0.000 | -0.014, 0.014 | 0.007 | 0.99 | -0.003 | -0.017, 0.01 | -0.475 | 0.63 |
| Metabolic dysfunction-associated fatty liver disease | 0.009 | -0.005, 0.022 | 1.207 | 0.23 | -0.004 | -0.018, 0.009 | -0.617 | 0.54 |
| Metabolic dysfunction-associated steatotic liver disease | 0.001 | -0.012, 0.015 | 0.207 | 0.84 | -0.005 | -0.019, 0.008 | -0.778 | 0.44 |
| **White matter hyperintensities**[2] | | | | | | | | |
| White matter hyperintensities | 0.002 | -0.012, 0.016 | 0.268 | 0.79 | -0.006 | -0.02, 0.008 | -0.880 | 0.38 |
| **Symbol digit substitution** | | | | | Tower rearranging | | | |
| **Cardiometabolic risk factors**[1] | | | | | | | | |
| Body mass index | -0.017 | -0.03, -0.003 | -2.385 | 0.017 | -0.001 | -0.015, 0.013 | -0.139 | 0.89 |
| Waist circumference | -0.009 | -0.023, 0.004 | -1.330 | 0.18 | 0.002 | -0.012, 0.016 | 0.290 | 0.77 |
| Systolic blood pressure | 0.004 | -0.009, 0.018 | 0.600 | 0.55 | -0.001 | -0.015, 0.013 | -0.151 | 0.88 |
| Diastolic blood pressure | 0.001 | -0.012, 0.015 | 0.176 | 0.86 | 0.004 | -0.01, 0.018 | 0.580 | 0.56 |
| Pulse pressure | 0.006 | -0.007, 0.02 | 0.896 | 0.37 | -0.006 | -0.019, 0.008 | -0.819 | 0.41 |
| C-reactive protein | 0.000 | -0.014, 0.013 | -0.040 | 0.97 | 0.003 | -0.01, 0.017 | 0.474 | 0.64 |
| Glycated hemoglobin | -0.004 | -0.018, 0.009 | -0.625 | 0.53 | 0.004 | -0.009, 0.018 | 0.613 | 0.54 |
| High-density lipoprotein cholesterol | 0.005 | -0.009, 0.018 | 0.660 | 0.51 | -0.001 | -0.014, 0.013 | -0.089 | 0.93 |
| Low-density lipoprotein cholesterol | 0.011 | -0.003, 0.025 | 1.557 | 0.12 | 0.010 | -0.004, 0.024 | 1.429 | 0.15 |
| Total cholesterol | 0.010 | -0.003, 0.024 | 1.480 | 0.14 | 0.011 | -0.003, 0.024 | 1.499 | 0.13 |
| Triglycerides | 0.005 | -0.008, 0.019 | 0.750 | 0.45 | -0.001 | -0.014, 0.013 | -0.098 | 0.92 |
| **Cardiometabolic principal components**[1] | | | | | | | | |
| Principal component 1 | -0.001 | -0.015, 0.012 | -0.179 | 0.86 | 0.001 | -0.012, 0.015 | 0.208 | 0.84 |
| Principal component 2 | -0.010 | -0.024, 0.004 | -1.457 | 0.15 | -0.011 | -0.025, 0.003 | -1.568 | 0.12 |
| Principal component 3 | 0.004 | -0.01, 0.017 | 0.512 | 0.61 | -0.005 | -0.019, 0.008 | -0.761 | 0.45 |

| | Partial correlation coefficient | 95% confidence interval | T-statistic | P-value | Partial correlation coefficient | 95% confidence interval | T-statistic | P-value |
|---|---|---|---|---|---|---|---|---|
| **Liver fat and steatotic liver disease[1]** | | | | | | | | |
| Liver fat | -0.002 | -0.016, 0.011 | -0.345 | 0.73 | 0.011 | -0.003, 0.025 | 1.541 | 0.12 |
| Nonalcoholic fatty liver disease | 0.002 | -0.012, 0.015 | 0.227 | 0.82 | 0.005 | -0.009, 0.019 | 0.740 | 0.46 |
| Metabolic dysfunction-associated fatty liver disease | 0.001 | -0.012, 0.015 | 0.172 | 0.86 | 0.002 | -0.012, 0.015 | 0.241 | 0.81 |
| Metabolic dysfunction-associated steatotic liver disease | 0.002 | -0.012, 0.015 | 0.220 | 0.83 | 0.005 | -0.009, 0.018 | 0.650 | 0.52 |
| **White matter hyperintensities[2]** | | | | | | | | |
| White matter hyperintensities | -0.006 | -0.019, 0.008 | -0.820 | 0.41 | 0.000 | -0.013, 0.014 | 0.067 | 0.95 |
| **Paired associate learning** | | | | | **Pairs matching** | | | |
| **Cardiometabolic risk factors[2]** | | | | | | | | |
| Body mass index | -0.016 | -0.029, -0.004 | -2.530 | 0.011 | 0.003 | -0.009, 0.014 | 0.447 | 0.65 |
| Waist circumference | -0.003 | -0.016, 0.009 | -0.513 | 0.61 | 0.005 | -0.007, 0.016 | 0.816 | 0.41 |
| Systolic blood pressure | -0.007 | -0.02, 0.006 | -1.038 | 0.3 | -0.005 | -0.016, 0.006 | -0.889 | 0.37 |
| Diastolic blood pressure | -0.011 | -0.023, 0.002 | -1.617 | 0.11 | -0.003 | -0.014, 0.008 | -0.538 | 0.59 |
| Pulse pressure | 0.000 | -0.013, 0.013 | 0.033 | 0.97 | -0.004 | -0.015, 0.007 | -0.751 | 0.45 |
| C-reactive protein | -0.002 | -0.015, 0.01 | -0.374 | 0.71 | -0.002 | -0.013, 0.009 | -0.404 | 0.69 |
| Glycated hemoglobin | 0.007 | -0.006, 0.02 | 1.094 | 0.27 | 0.014 | 0.003, 0.025 | 2.415 | 0.016 |
| High-density lipoprotein cholesterol | -0.001 | -0.014, 0.011 | -0.194 | 0.85 | 0.004 | -0.008, 0.015 | 0.640 | 0.52 |
| Low-density lipoprotein cholesterol | 0.007 | -0.006, 0.02 | 1.100 | 0.27 | -0.002 | -0.013, 0.01 | -0.284 | 0.78 |
| Total cholesterol | 0.005 | -0.007, 0.018 | 0.831 | 0.41 | 0.000 | -0.012, 0.011 | -0.062 | 0.95 |
| Triglycerides | 0.010 | -0.002, 0.023 | 1.599 | 0.11 | 0.005 | -0.007, 0.016 | 0.789 | 0.43 |
| **Cardiometabolic principal components[1]** | | | | | | | | |
| Principal component 1 | -0.007 | -0.02, 0.006 | -1.081 | 0.28 | -0.003 | -0.015, 0.008 | -0.607 | 0.54 |
| Principal component 2 | -0.003 | -0.016, 0.01 | -0.491 | 0.62 | 0.003 | -0.008, 0.014 | 0.562 | 0.57 |
| Principal component 3 | -0.005 | -0.018, 0.008 | -0.752 | 0.45 | -0.003 | -0.014, 0.008 | -0.551 | 0.58 |
| **Liver fat and steatotic liver disease[1]** | | | | | | | | |
| Liver fat | 0.010 | -0.003, 0.023 | 1.516 | 0.13 | 0.003 | -0.008, 0.015 | 0.589 | 0.56 |
| Nonalcoholic fatty liver disease | 0.010 | -0.003, 0.023 | 1.522 | 0.13 | 0.004 | -0.007, 0.015 | 0.730 | 0.47 |
| Metabolic dysfunction-associated fatty liver disease | 0.002 | -0.01, 0.015 | 0.358 | 0.72 | 0.008 | -0.003, 0.019 | 1.419 | 0.16 |
| Metabolic dysfunction-associated steatotic liver disease | 0.010 | -0.003, 0.022 | 1.498 | 0.13 | 0.004 | -0.007, 0.015 | 0.749 | 0.45 |
| **White matter hyperintensities[2]** | | | | | | | | |
| White matter hyperintensities | -0.008 | -0.021, 0.005 | -1.235 | 0.22 | 0.002 | -0.009, 0.013 | 0.297 | 0.77 |

*Notes:* Interaction effects derived from multiple linear regression analyses between predictors (each predictor analyzed in a separate regression model) and sex (predictor-by-sex) on the outcome variables (the individual cognitive tests).

[1]Adjusted for age, age$^2$, sex, age-by-sex, age$^2$-by-sex, assessment center, smoking status, alcohol consumption, and education.

[2]Adjusted for age, age$^2$, sex, age-by-sex, age$^2$-by-sex, assessment center, smoking status, alcohol consumption, education, and intracranial volume.

**Table S15: Mediation analyses between cardiometabolic factors and white matter hyperintensities, with liver fat as mediator**

| | Direct effect | | | Indirect effect | | | Total effect | | |
|---|---|---|---|---|---|---|---|---|---|
| **Predictor** | **Beta std.** | **Z-score** | **P-value** | **Beta std.** | **Z-score** | **P-value** | **Beta std.** | **Z-score** | **P-value** |
| **Cardiometabolic risk factors[1]** | | | | | | | | | |
| Body mass index | 0.09 | 16.57 | 0 | 0.02 | 9.21 | 0 | 0.11 | 22.09 | 0 |
| Waist circumference | 0.10 | 16.67 | 0 | 0.02 | 9.10 | 0 | 0.12 | 22.21 | 0 |
| Systolic blood pressure | 0.10 | 18.63 | 0 | 0.01 | 12.56 | 0 | 0.11 | 20.72 | 0 |
| Diastolic blood pressure | 0.10 | 20.72 | 0 | 0.01 | 12.37 | 0 | 0.11 | 23.59 | 0 |
| Pulse pressure | 0.05 | 9.46 | 0 | 0.00 | 5.83 | $5.7x10^{-9}$ | 0.05 | 10.04 | 0 |
| C-reactive protein | 0.03 | 5.16 | $2.5x10^{-7}$ | 0.02 | 14.45 | 0 | 0.05 | 9.29 | 0 |
| Glycated hemoglobin | 0.04 | 7.29 | $3.1x10^{-13}$ | 0.01 | 12.88 | 0 | 0.05 | 9.74 | 0 |
| High-density lipoprotein cholesterol | -0.02 | -3.87 | 0.0001 | -0.02 | -15.13 | 0 | -0.04 | -8.38 | 0 |
| Low-density lipoprotein cholesterol | 0.00 | -0.43 | 0.67 | 0.01 | 11.51 | 0 | 0.01 | 1.10 | 0.27 |
| Total cholesterol | 0.00 | -0.88 | 0.38 | 0.00 | 7.26 | $3.9x10^{-13}$ | 0.00 | -0.09 | 0.93 |
| Triglycerides | 0.01 | 2.82 | 0.0048 | 0.03 | 14.94 | 0 | 0.04 | 8.40 | 0 |
| **Cardiometabolic principal components[1]** | | | | | | | | | |
| Principal component 1 | 0.12 | 22.02 | 0 | 0.02 | 8.17 | $2.2x10^{-16}$ | 0.14 | 26.98 | 0 |
| Principal component 2 | 0.02 | 4.89 | $1x10^{-6}$ | 0.01 | 13.06 | 0 | 0.03 | 6.91 | $4.9x10^{-12}$ |
| Principal component 3 | 0.02 | 4.88 | $1x10^{-6}$ | -0.02 | -16.55 | 0 | 0.00 | 0.83 | 0.4 |

*Notes:* The direct, indirect (via the mediator liver fat), and total effects of the predictors (each predictor analyzed in a separate model) on white matter hyperintensities derived from structural equation modeling mediation models. Beta std., standardized regression coefficient.
[1]Adjusted for age, $age^2$, sex, assessment center, smoking status, and alcohol consumption.

**Table S16: Sex-stratified mediation analyses between cardiometabolic factors and white matter hyperintensities, with liver fat as mediator**

| | Male | | | Female | | |
|---|---|---|---|---|---|---|
| | **Beta std.** | **Z-score** | **P-value** | **Beta std.** | **Z-score** | **P-value** |
| **Body mass index** | | | | | | |
| **Direct effect** | 0.12 | 23.29 | 0 | 0.07 | 9.11 | 0 |
| **Indirect effect** | 0.02 | 8.17 | $2.2x10^{-16}$ | 0.02 | 5.81 | $6.4x10^{-9}$ |
| **Total effect** | 0.14 | 25.23 | 0 | 0.08 | 12.74 | 0 |
| **Waist circumference** | | | | | | |
| **Direct effect** | 0.11 | 15.55 | 0 | 0.07 | 9.27 | 0 |
| **Indirect effect** | 0.02 | 6.18 | $6.6x10^{-10}$ | 0.02 | 5.49 | $3.9x10^{-8}$ |
| **Total effect** | 0.13 | 18.78 | 0 | 0.08 | 12.91 | 0 |
| **Diastolic blood pressure** | | | | | | |
| **Direct effect** | 0.09 | 9.51 | 0 | 0.11 | 17.22 | 0 |
| **Indirect effect** | 0.01 | 8.88 | 0 | 0.01 | 6.80 | $1.1x10^{-11}$ |
| **Total effect** | 0.10 | 11.82 | 0 | 0.12 | 19.07 | 0 |
| **C-reactive protein** | | | | | | |
| **Direct effect** | 0.03 | 3.43 | $6x10^{-4}$ | 0.02 | 3.19 | 0.0014 |
| **Indirect effect** | 0.02 | 15.18 | 0 | 0.02 | 8.92 | 0 |
| **Total effect** | 0.05 | 5.07 | $4x10^{-7}$ | 0.04 | 6.30 | $2.9x10^{-10}$ |
| **Low-density lipoprotein cholesterol** | | | | | | |
| **Direct effect** | -0.02 | -2.34 | 0.019 | 0.00 | 0.49 | 0.62 |
| **Indirect effect** | 0.00 | 5.91 | $3.4x10^{-9}$ | 0.01 | 8.47 | 0 |
| **Total effect** | -0.01 | -1.74 | 0.081 | 0.01 | 1.67 | 0.095 |
| **Triglycerides** | | | | | | |
| **Direct effect** | 0.01 | 1.45 | 0.15 | 0.01 | 1.91 | 0.056 |
| **Indirect effect** | 0.03 | 16.19 | 0 | 0.02 | 9.08 | 0 |
| **Total effect** | 0.03 | 6.18 | $6.6x10^{-10}$ | 0.04 | 5.79 | $7.2x10^{-9}$ |
| **Principal component 2** | | | | | | |
| **Direct** | 0.05 | 5.00 | $5.9x10^{-7}$ | 0.01 | 1.70 | 0.089 |
| **Indirect effect** | 0.01 | 7.78 | $7.5x10^{-15}$ | 0.01 | 8.84 | 0 |
| **Total effect** | 0.06 | 6.34 | $2.3x10^{-10}$ | 0.02 | 3.19 | 0.0014 |

*Notes:* The direct, indirect (via the mediator liver fat), and total effects of the predictors (each predictor analyzed in a separate model) on white matter hyperintensities derived from structural equation modeling mediation models stratified by sex. The models are adjusted for age, age$^2$, assessment center, smoking status, and alcohol consumption. Beta std., standardized regression coefficient.

### Table S17: Mediation analyses between liver fat and the cognitive principal component 1, with WMH as mediator

| | **Direct effect** | | | **Indirect effect** | | | **Total effect** | | |
|---|---|---|---|---|---|---|---|---|---|
| **Predictor** | **Beta std.** | **Z-score** | **P-value** | **Beta std.** | **Z-score** | **P-value** | **Beta std.** | **Z-score** | **P-value** |
| Liver fat | -0.02 | -3.36 | 0.00078 | -0.01 | -8.38 | 0 | -0.03 | -4.42 | $1x10^{-5}$ |
| Nonalcoholic fatty liver disease | -0.02 | -3.47 | 0.00053 | 0.00 | -7.56 | $4.0x10^{-14}$ | -0.03 | -4.23 | $2.4x10^{-5}$ |
| Metabolic dysfunction-associated fatty liver disease | -0.02 | -3.59 | 0.00033 | -0.01 | -8.13 | $4.4x10^{-16}$ | -0.03 | -4.48 | $7.3x10^{-6}$ |
| Metabolic dysfunction-associated steatotic liver disease | -0.02 | -3.37 | 0.00075 | 0.00 | -7.60 | $2.9x10^{-14}$ | -0.02 | -4.14 | $3.4x10^{-5}$ |

*Notes:* The direct, indirect (via the mediator white matter hyperintensities), and total effects of the predictors (each predictor analyzed in a separate model) on the cognitive principal component 1 derived from structural equation modeling mediation models. The models are adjusted for age, age$^2$, sex, assessment center, smoking status, alcohol consumption, education, and intracranial volume. Beta std., standardized regression coefficient

**Table S18: Mediation analyses between liver fat and the individual cognitive tests, with WMH as mediator**

| | Direct effect | | | Indirect effect | | | Total effect | | |
|---|---|---|---|---|---|---|---|---|---|
| **Predictor** | **Beta std.** | **Z-score** | **P-value** | **Beta std.** | **Z-score** | **P-value** | **Beta std.** | **Z-score** | **P-value** |
| **Numeric memory** | | | | | | | | | |
| **Liver fat** | -0.038 | -5.999 | $2x10^{-9}$ | -0.003 | -4.928 | $8.3x10^{-7}$ | -0.042 | -6.562 | $5.3x10^{-11}$ |
| **Nonalcoholic fatty liver disease** | -0.010 | -1.748 | 0.08 | -0.004 | -6.716 | $1.9x10^{-11}$ | -0.014 | -2.546 | 0.011 |
| **Metabolic dysfunction-associated fatty liver disease** | -0.004 | -0.661 | 0.51 | 0.005 | 6.522 | $6.9x10^{-11}$ | 0.001 | 0.101 | 0.92 |
| **Metabolic dysfunction-associated steatotic liver disease** | -0.019 | -2.820 | 0.0048 | -0.005 | -6.196 | $5.8x10^{-10}$ | -0.024 | -3.526 | 0.00042 |
| **Fluid intelligence** | | | | | | | | | |
| **Liver fat** | -0.021 | -3.210 | 0.0013 | -0.005 | -7.083 | $1.4x10^{-12}$ | -0.026 | -4.067 | $4.8x10^{-5}$ |
| **Nonalcoholic fatty liver disease** | 0.029 | 4.223 | $2.4x10^{-5}$ | -0.004 | -4.858 | $1.2x10^{-6}$ | 0.026 | 3.713 | $2x10^{-4}$ |
| **Metabolic dysfunction-associated fatty liver disease** | -0.037 | -5.782 | $7.4x10^{-9}$ | -0.004 | -6.042 | $1.5x10^{-9}$ | -0.041 | -6.466 | $1x10^{-10}$ |
| **Metabolic dysfunction-associated steatotic liver disease** | -0.021 | -3.611 | 0.00031 | 0.002 | 3.293 | 0.00099 | -0.019 | -3.287 | 0.001 |
| **Trail making test B** | | | | | | | | | |
| **Liver fat** | -0.031 | -4.722 | $2.3x10^{-6}$ | -0.003 | -4.957 | $7.1x10^{-7}$ | -0.033 | -5.119 | $3.1x10^{-7}$ |
| **Nonalcoholic fatty liver disease** | -0.003 | -0.462 | 0.64 | -0.003 | -6.352 | $2.1x10^{-10}$ | -0.006 | -1.018 | 0.31 |
| **Metabolic dysfunction-associated fatty liver disease** | -0.001 | -0.110 | 0.91 | 0.004 | 6.012 | $1.8x10^{-9}$ | 0.003 | 0.430 | 0.67 |
| **Metabolic dysfunction-associated steatotic liver disease** | -0.022 | -3.256 | 0.0011 | -0.003 | -5.823 | $5.8x10^{-9}$ | -0.025 | -3.759 | 0.00017 |
| **Matrix test** | | | | | | | | | |
| **Liver fat** | -0.024 | -3.658 | 0.00025 | -0.004 | -6.553 | $5.6x10^{-11}$ | -0.028 | -4.266 | $2x10^{-5}$ |
| **Nonalcoholic fatty liver disease** | 0.018 | 2.600 | 0.0093 | -0.002 | -4.456 | $8.3x10^{-6}$ | 0.016 | 2.244 | 0.025 |
| **Metabolic dysfunction-associated fatty liver disease** | -0.033 | -5.167 | $2.4x10^{-7}$ | -0.003 | -5.807 | $6.4x10^{-9}$ | -0.036 | -5.663 | $1.5x10^{-8}$ |
| **Metabolic dysfunction-associated steatotic liver disease** | -0.018 | -3.148 | 0.0016 | 0.001 | 3.077 | 0.0021 | -0.017 | -2.916 | 0.0035 |
| **Symbol digit substitution** | | | | | | | | | |
| **Liver fat** | -0.034 | -5.296 | $1.2x10^{-7}$ | -0.003 | -4.987 | $6.1x10^{-7}$ | -0.037 | -5.766 | $8.1x10^{-9}$ |
| **Nonalcoholic fatty liver disease** | -0.005 | -0.985 | 0.32 | -0.004 | -6.490 | $8.6x10^{-11}$ | -0.009 | -1.636 | 0.1 |
| **Metabolic dysfunction-associated fatty liver disease** | 0.000 | 0.050 | 0.96 | 0.004 | 6.285 | $3.3x10^{-10}$ | 0.004 | 0.671 | 0.5 |
| **Metabolic dysfunction-associated steatotic liver disease** | -0.016 | -2.372 | 0.018 | -0.004 | -6.061 | $1.3x10^{-9}$ | -0.020 | -2.951 | 0.0032 |
| **Tower rearranging** | | | | | | | | | |
| **Liver fat** | -0.025 | -3.960 | $7.5x10^{-5}$ | -0.004 | -6.895 | $5.4x10^{-12}$ | -0.030 | -4.660 | $3.2x10^{-6}$ |
| **Nonalcoholic fatty liver disease** | 0.026 | 3.709 | 0.00021 | -0.003 | -4.628 | $3.7x10^{-6}$ | 0.023 | 3.294 | 0.00099 |
| **Metabolic dysfunction-associated fatty liver disease** | -0.038 | -5.979 | $2.2x10^{-9}$ | -0.004 | -5.820 | $5.9x10^{-9}$ | -0.042 | -6.548 | $5.8x10^{-11}$ |
| **Metabolic dysfunction-associated steatotic liver disease** | -0.022 | -3.874 | 0.00011 | 0.002 | 3.237 | 0.0012 | -0.021 | -3.601 | 0.00032 |
| **Paired associate learning** | | | | | | | | | |
| **Liver fat** | -0.030 | -4.658 | $3.2x10^{-6}$ | -0.003 | -4.876 | $1.1x10^{-6}$ | -0.033 | -5.074 | $3.9x10^{-7}$ |
| **Nonalcoholic fatty liver disease** | -0.002 | -0.297 | 0.77 | -0.003 | -6.422 | $1.3x10^{-10}$ | -0.005 | -0.853 | 0.39 |
| **Metabolic dysfunction-associated fatty liver disease** | 0.001 | 0.096 | 0.92 | 0.004 | 5.930 | $3x10^{-9}$ | 0.004 | 0.636 | 0.52 |
| **Metabolic dysfunction-associated steatotic liver disease** | -0.022 | -3.218 | 0.0013 | -0.003 | -5.854 | $4.8x10^{-9}$ | -0.025 | -3.729 | 0.00019 |
| **Pairs matching** | | | | | | | | | |
| **Liver fat** | -0.024 | -3.642 | 0.00027 | -0.004 | -6.570 | $5x10^{-11}$ | -0.028 | -4.268 | $2x10^{-5}$ |
| **Nonalcoholic fatty liver disease** | 0.019 | 2.770 | 0.0056 | -0.003 | -4.446 | $8.7x10^{-6}$ | 0.017 | 2.413 | 0.016 |
| **Metabolic dysfunction-associated fatty liver disease** | -0.034 | -5.283 | $1.3x10^{-7}$ | -0.003 | -5.799 | $6.7x10^{-9}$ | -0.038 | -5.772 | $7.8x10^{-9}$ |
| **Metabolic dysfunction-associated steatotic liver disease** | -0.021 | -3.534 | 0.00041 | 0.001 | 3.166 | 0.0015 | -0.019 | -3.304 | 0.00095 |

*Notes:* The direct, indirect (via the mediator white matter hyperintensities), and total effects of the predictors (each predictor analyzed in a separate model) on the outcomes (the individual cognitive tests) derived from structural equation modeling mediation models. The models are adjusted for age, $age^2$, sex, assessment center, smoking status, alcohol consumption, education, and intracranial volume. Beta std., standardized regression coefficient

### Supplementary Figures

**Figure S1: Correlations of cardiometabolic risk factors**

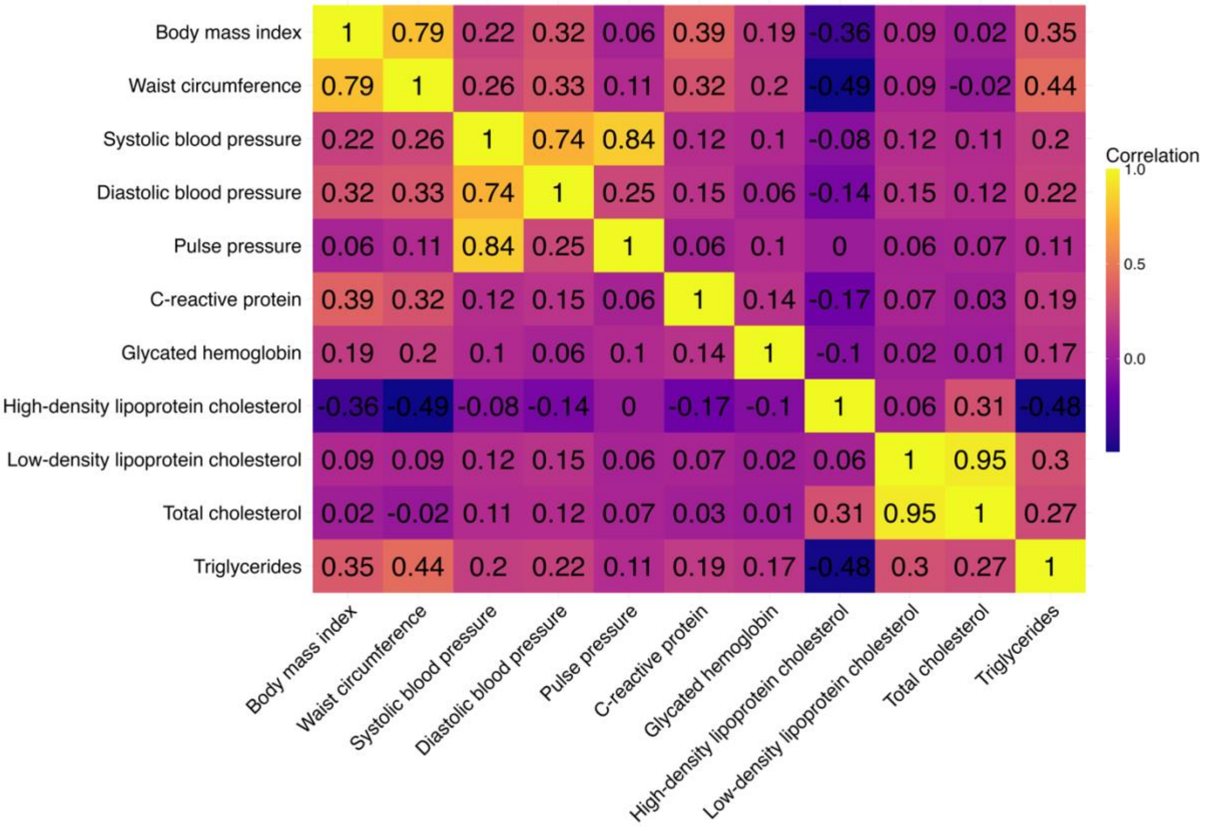

*Notes:* Pearson cross-correlation matrix of the 11 included cardiometabolic tests.

**Figure S2: Histograms of cardiometabolic and imaging variables**

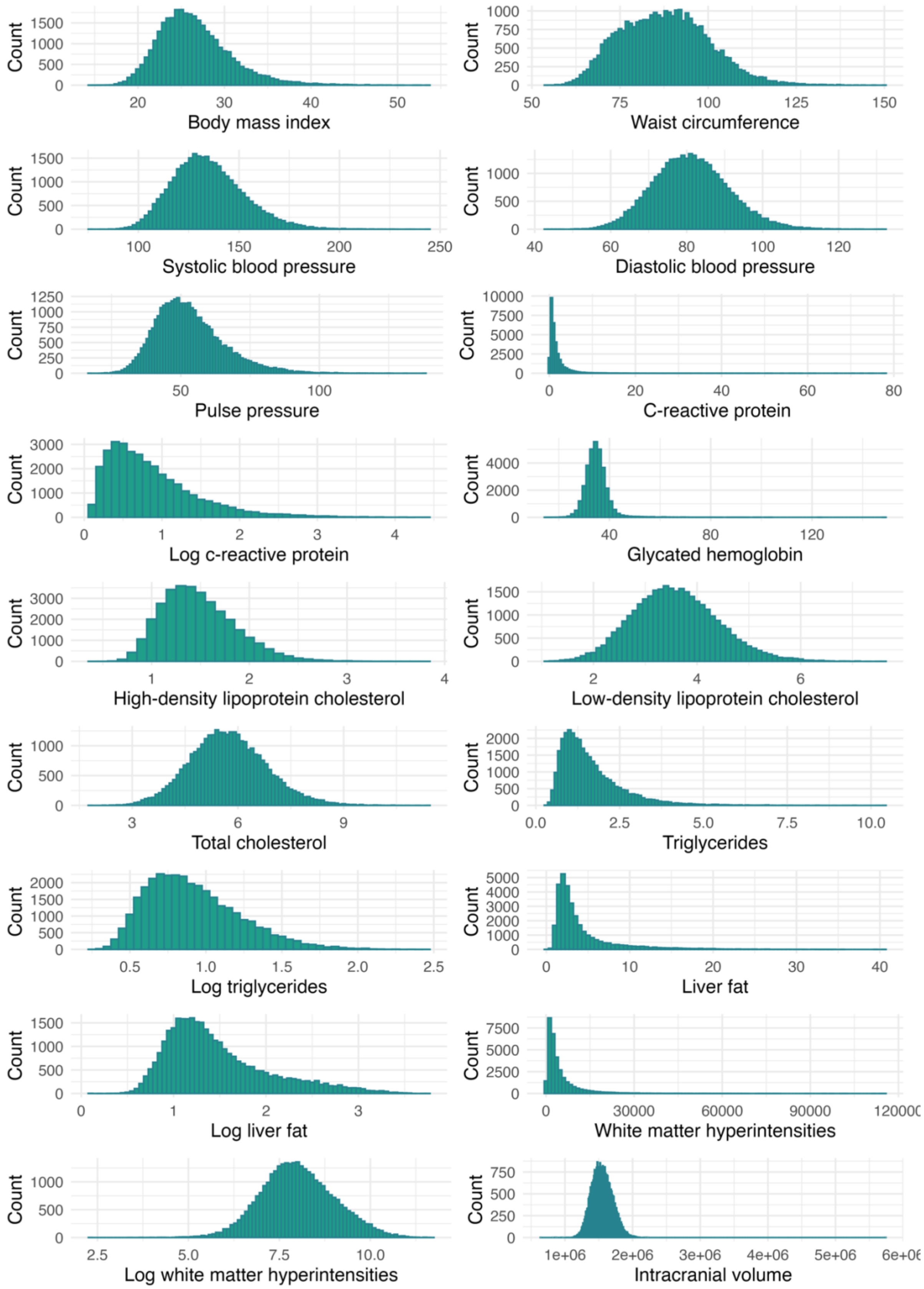

**Figure S3: Quantile-quantile plots of cardiometabolic and imaging variables**

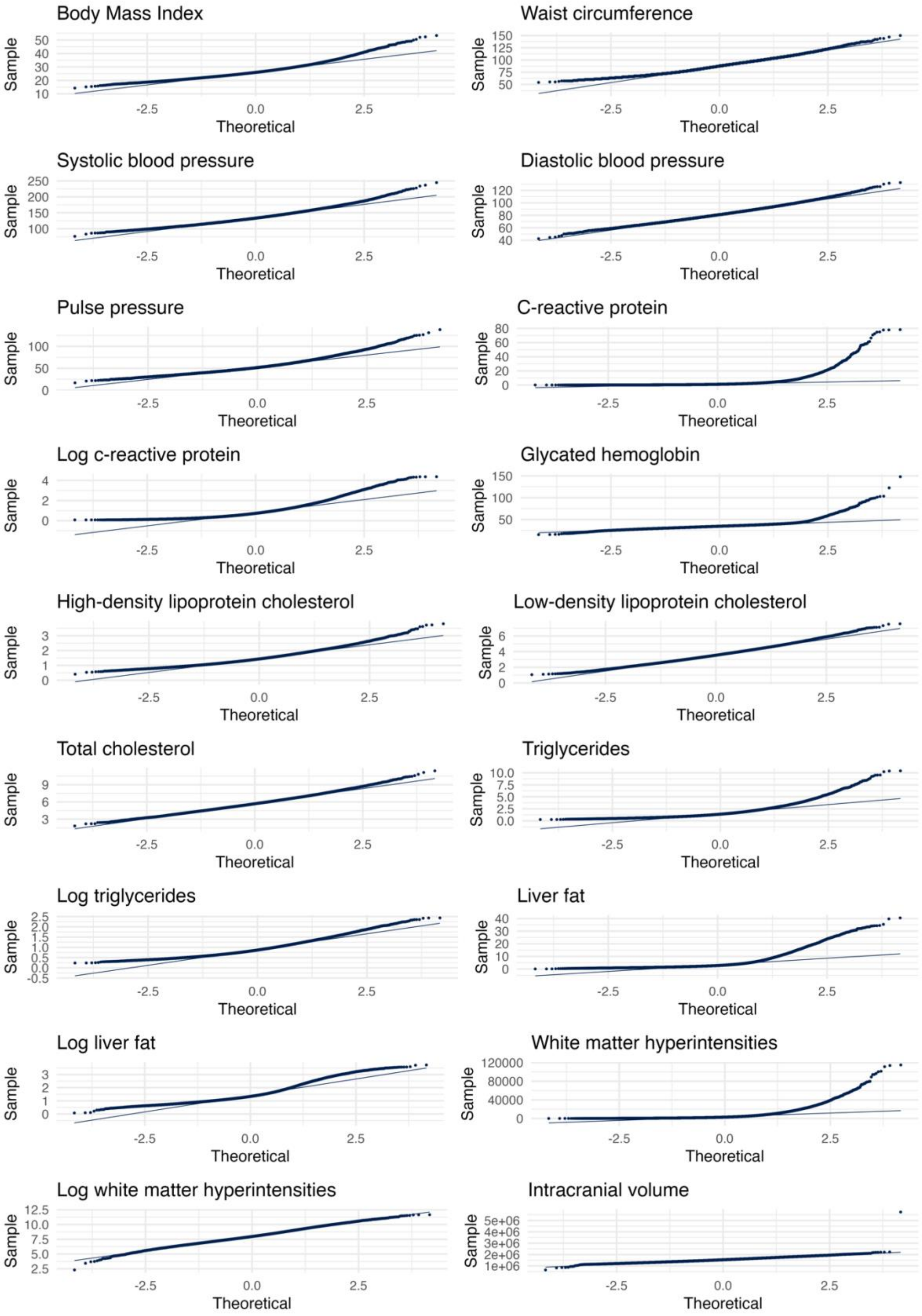

*Notes:* Y-axes, sample quantiles; x-axes theoretical quantiles.

**Figure S4: Scree plot cardiometabolic principal component analysis**

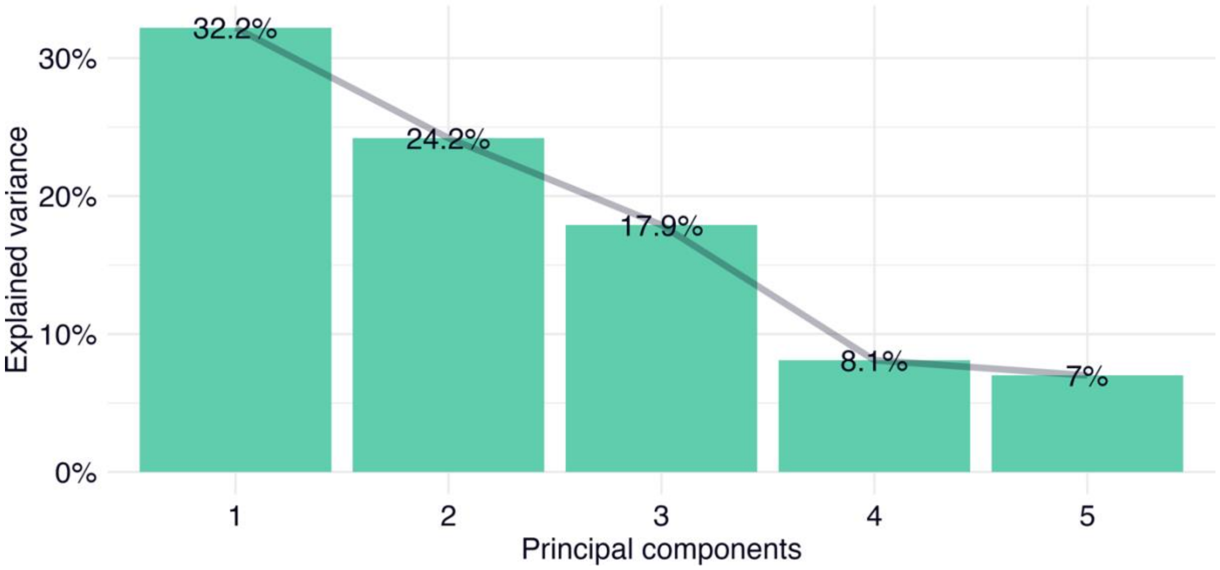

*Notes:* Scree plot of the explained variance of the included 11 cardiometabolic variables for the cardiometabolic principal components 1-5.

**Figure S5: Loadings cardiometabolic principal component analysis**

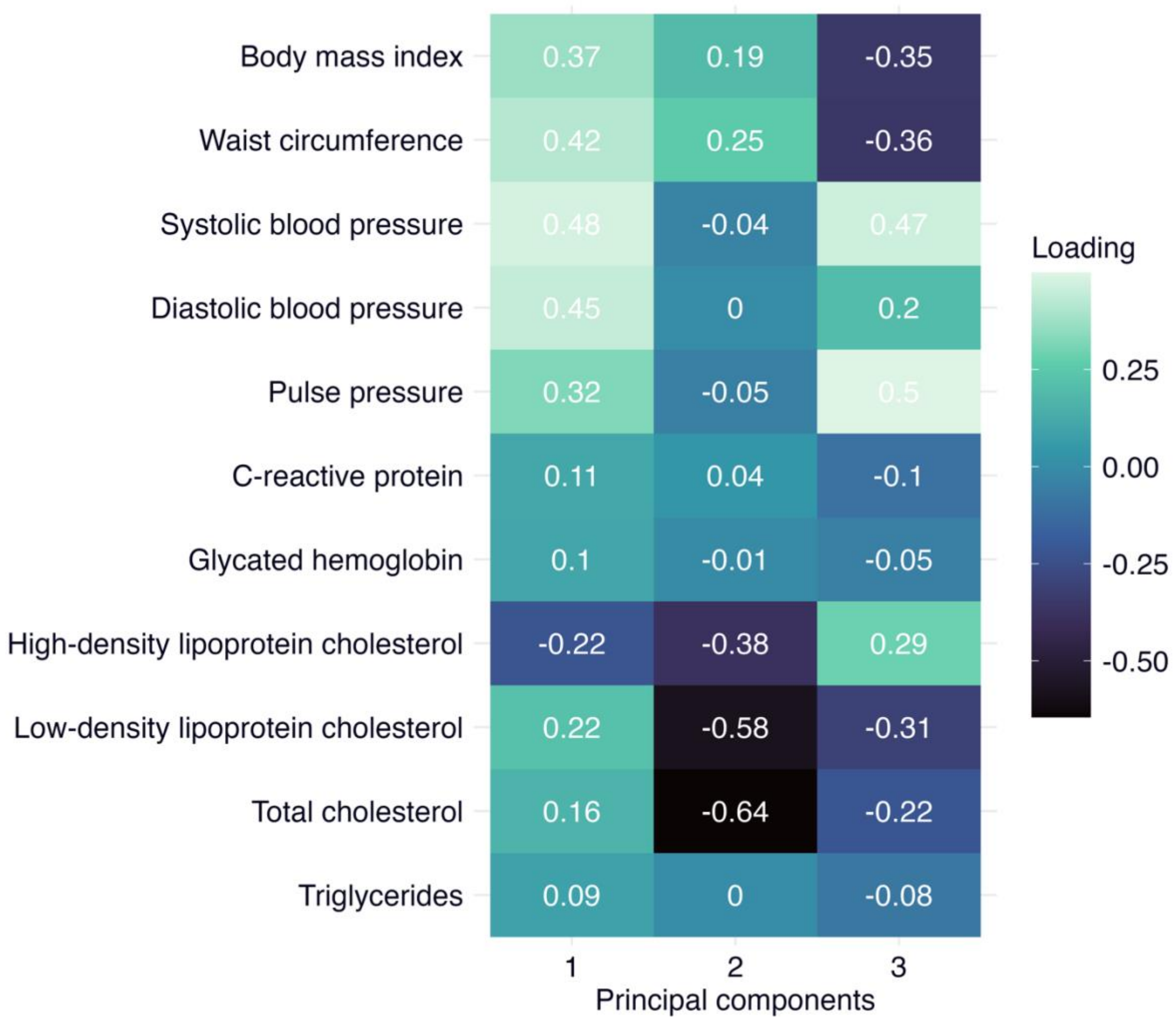

*Notes:* Heat map of the loadings from the 11 included cardiometabolic variables onto the cardiometabolic principal components 1-3.

**Figure S6: Correlations of cognitive variables**

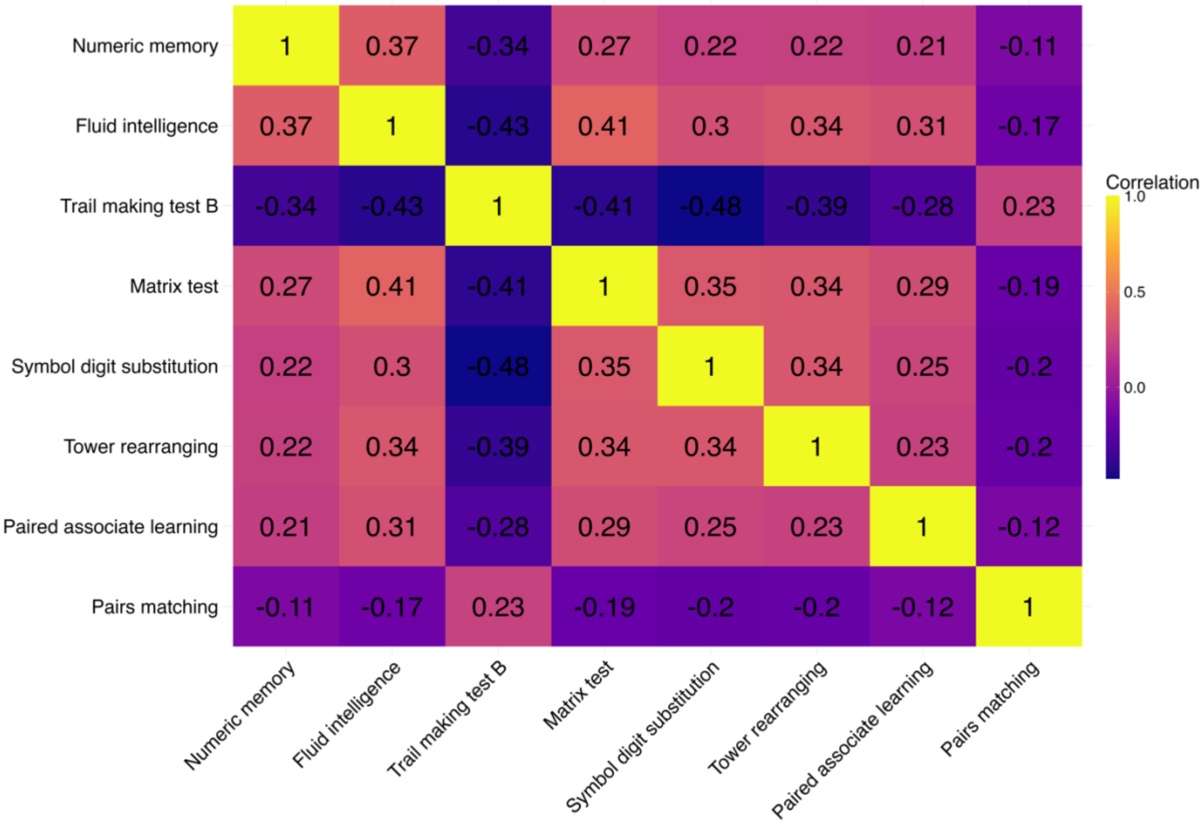

*Notes:* Pearson cross-correlation matrix of the 8 included cognitive tests.

**Figure S7: Histograms of cognitive tests**

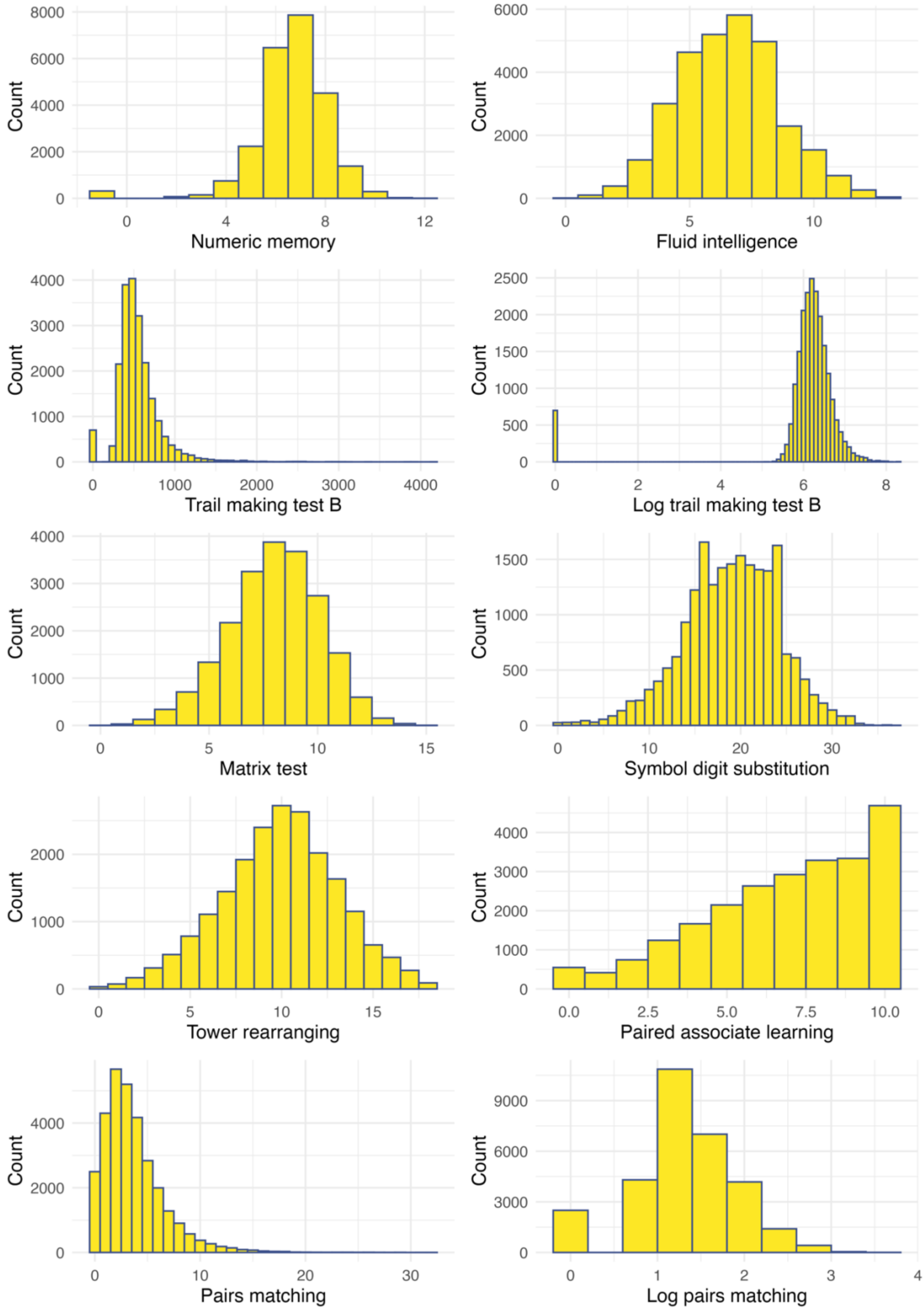

**Figure S8: Quantile-quantile plots of cognitive tests**

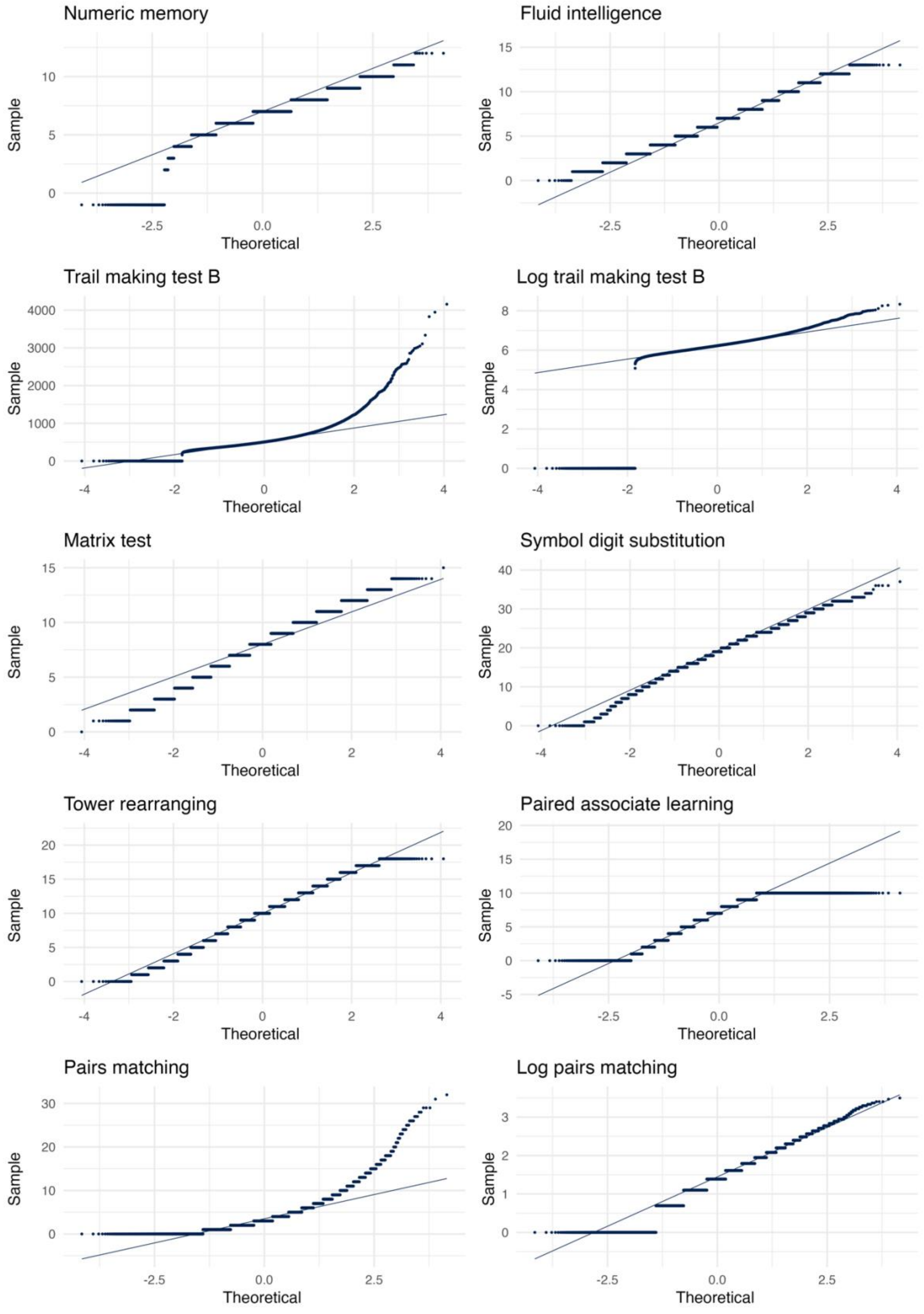

Notes: Y-axes, sample quantiles; x-axes theoretical quantiles.

**Figure S9: Scree plot cognitive performance principal component analysis**

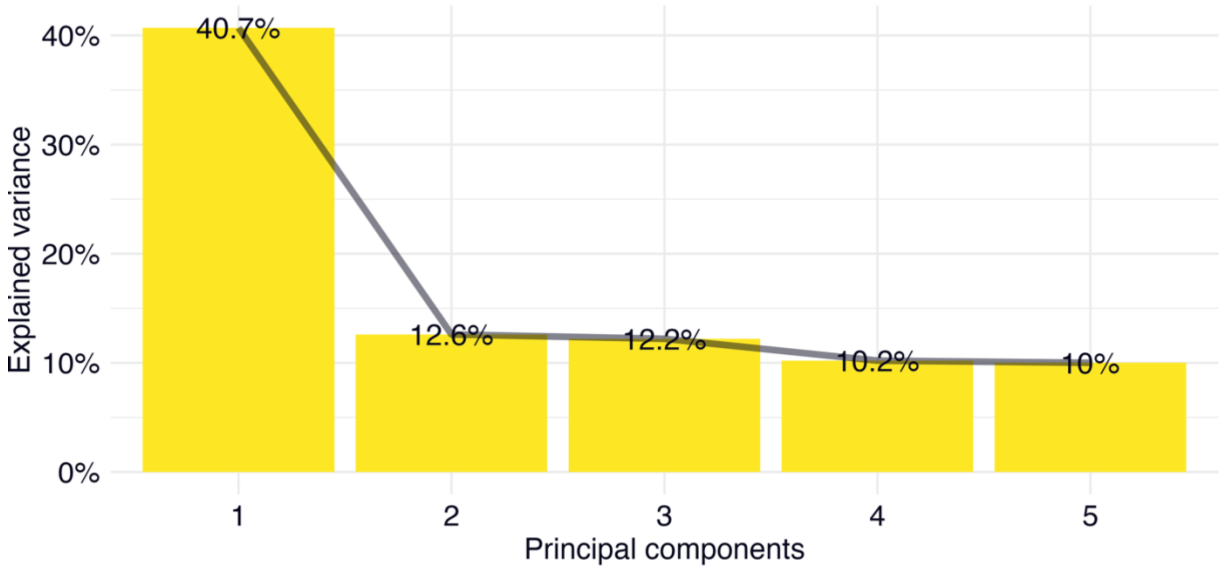

*Notes:* Scree plot of the explained variance of the included 8 cognitive tests for the cognitive principal components 1-5.

**Figure S10: Loadings of cognitive tests onto cognitive principal component 1**

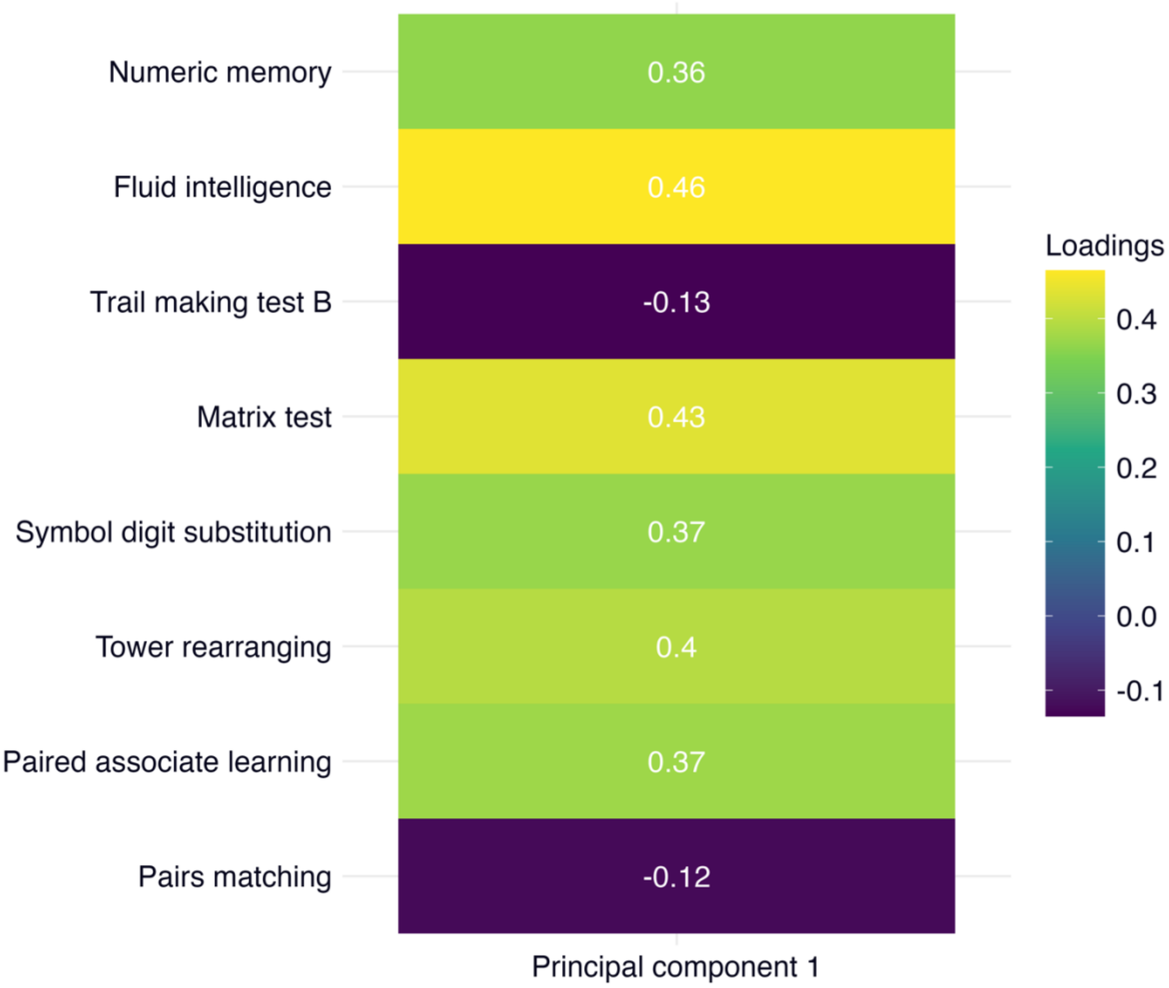

*Notes:* Heat map of the loadings from the included 8 cognitive tests onto the cognitive principal component 1.
